# Polygenic Susceptibility to Hypertension is Associated with Worse Cognitive Performance in Middle-Aged Persons without Dementia

**DOI:** 10.1101/2022.09.27.22280434

**Authors:** Cyprien Rivier, Natalia Szejko, Daniela Renedo, Rommell Noche, Julian N. Acosta, Cameron P. Both, Seyedmehdi Payabvash, Adam De Havenon, Kevin N. Sheth, Thomas M. Gill, Guido J. Falcone

## Abstract

**Background:** Mounting evidence indicates that hypertension leads to higher risk of cognitive decline and dementia. Hypertension is a highly heritable trait and a higher polygenic susceptibility to hypertension (PSH) is known to be associated with higher risk of dementia. We tested the hypothesis that a higher PSH leads to worse cognitive performance in middle-aged persons without dementia.

**Methods:** We conducted a nested, cross-sectional, genetic study within the UK Biobank, a large population study that enrolled middle-aged Britons. Study participants with a history of dementia or stroke were excluded. We categorized participants as having low (≤20^th^ percentile), intermediate (>20^th^ and <80^th^ percentile), or high (≥80^th^ percentile) PSH according to results of 2 polygenic risk scores for systolic and diastolic blood pressure (BP), generated with genomic data on 732 genetic risk variants for these traits. Cognitive performance was evaluated via 5 simple tests: Pairs Memory, Reaction Time, Numeric Memory, Prospective Memory and Fluid Intelligence. A general cognitive ability score was calculated as the first principal component of a principal component analysis that included the results of these 5 tests. Primary analyses focused on Europeans and secondary analyses included all race/ethnic groups.

**Results:** Out of 409,551 study participants of European ancestry with available genomic data, 42,080 (10.3%) completed all 5 tests. Multivariable regression models using systolic BP-related genetic variants indicated that, compared to study participants with low PSH, those with intermediate and high PSH had reductions of 3.9% (beta -0.039, SE 0.012) and 6.6% (beta - 0.066, SE 0.014), respectively, in their general cognitive ability score (test for trend p <0.001). Secondary analyses including all race/ethnic groups (n=48,118) and using diastolic BP-related genetic variants yielded similar results (both instances, p<0.05). Analyses evaluating each cognitive test separately indicated that Reaction Time, Numeric Memory and Fluid Intelligence drove the association of PSH with the general cognitive ability score (all individual tests, p<0.05).

**Conclusions:** Among non-demented, community-dwelling, middle-aged Britons, a higher PSH is associated with worse cognitive performance. These findings suggest the genetic predisposition to hypertension influence brain health in persons who have not yet developed dementia.

## INTRODUCTION

Mounting evidence indicates that preserving brain health during middle age leads to substantial health benefits later in life.^1^ Cognitive decline and dementia constitute two of the most important contributors to poor brain health in older adults.^2^ Because hypertension is emerging as an important determinant of declining cognitive function, blood pressure reduction is actively being explored as a protective strategy. In the SPRINT MIND clinical trial, a systolic blood pressure goal of <120 mmHg, compared to <140 mmHg, did not reduce the risk of probable dementia but did decrease the risk of cognitive decline and the composite risk of probable dementia and cognitive decline.^3^ Along the same lines, an ancillary neuroimaging study of this clinical trial showed that this treatment goal for hypertension was associated with a slower progression of white matter hyperintensities,^4^ a well-established marker of silent cerebrovascular disease observed in brain magnetic resonance imaging.^5,6^

Blood pressure is a highly heritable trait, with recent analysis estimating that up to 31% of its variance can be explained by genetic variation.^7^ This substantial genetic contribution has allowed the identification of numerous genetic variants associated with elevated systolic and diastolic blood pressure (BP), and, ultimately, formally diagnosed hypertension.^8–10^ Of note, most of these genetic risk variants are frequent, appearing in 1 to 50% of the population. When considered in isolation, each of these susceptibility loci has a small effect on observed BP; however, the aggregate contribution of all known genetic risk variants for elevated BP and hypertension can have a large effect on observed blood pressure.^11^ This polygenic susceptibility to hypertension (PSH) can be modeled via polygenic risk scores, a well-established tool in statistical genetics that calculates the aggregate contribution of numerous genetic risk variants for a disease or trait of interest.^12^

A higher PSH in middle age adults has been found to be associated with a higher risk of dementia of any cause later in life.^13^ Given these findings, the evaluation of the role of PSH in the cognitive performance of persons who have not yet developed dementia seems like an important and natural next step in this line of research. We hypothesize that a higher PSH, modeled through polygenic risk scores, is associated with worse cognitive performance in community-dwelling, middle-aged adults without a history of dementia or stroke.

## METHODS

### Study design and inclusion criteria

We conducted a nested study within the UK Biobank, a population-based study that enrolled ∼500,000 persons across the United Kingdom.^14^ At recruitment, participants provided electronically signed consent, answered questions on socio-demographic, lifestyle and health-related factors, completed a range of physical measures and cognitive tests (see below), and provided blood, urine and saliva samples that were stored for future testing. This large prospective study received approval by the North West Centre for Research Ethics Committee. All participants provided informed consent and agreed to follow up through linkage to their health-related records. In the present nested study, we included participants who completed the battery of cognitive tests and did not have a prior medical history of dementia or stroke.

### Genetic data

A detailed description of DNA collection and genotyping procedures is available elsewhere.^14^ Briefly, DNA was extracted from stored blood samples collected at the time of enrollment. Genotyping was carried out by the Affymetrix Research Services Laboratory in 106 sequential batches of approximately 4,700 samples each. A subset of 49,950 participants involved in the UK Biobank Lung Exome Variant Evaluation study were genotyped using the Affymetrix’s Applied Biosystems UK BiLEVE Axiom Array that contains 807,411 markers. Following this, 438,427 participants were genotyped using the closely related Applied Biosystems UK Biobank Axiom Array that contains 825,927 markers, of which 95% are shared with UK BiLEVE Axiom Array. Standard quality control procedures for genome-wide data were performed centrally by the UK Biobank research team, as previously reported,^14^ on markers that were present in both the UK BiLEVE Axiom Array and the UK Biobank Axiom Array. Genotype-level filters included testing for batch effects, plate effects, departures from Hardy-Weinberg equilibrium, sex effects, array effects, and discordance across control replicates. Subject-level quality checks included relatedness and heterozygosity. Principal component analysis was used to evaluate and assign ancestry.^16^ This pipeline yielded 488,377 samples and 805,426 markers that entered the imputation process, that was completed using the IMPUTE4 software^17^ and a combination of 3 reference panels: Haplotype Reference Consortium,^18^ UK10K haplotype^19^ and 1000 Genomes Phase 3.^20,21^ The result of the imputation process was a dataset with 93,095,623 autosomal SNPs, short indels and large structural variants in 487,442 individuals, including 409,551 of European ancestry and 77,891 belonged to other race/ethnic groups.

### Exposure ascertainment

Our exposure of interest was PSH modeled through polygenic risk scores, a well-established tool in statistical genetics that estimates an individual’s genetic burden across numerous genetic risk variants.^11^ For a given study participant, the polygenic risk score is the sum of the product of the risk allele counts for each variant multiplied by the allele’s reported effect on BP. To model the PSH, we built two separate polygenic risk scores, one for systolic and another for diastolic BP. Each of these scores used genetic information on 732 genetic risk variants known to be associated with higher systolic and diastolic BP (Supplemental Table 1). These genetic risk variants are single nucleotide polymorphisms with a minor allele frequency >1%, independent (r^2^, a measure of correlation between variants, <0.1), biallelic (involve two alleles only) and associated with one of the two BP traits of interest at genome-wide levels (p<5×10^−8^).^10^ Genetic variants included in the score were aligned to the GRCh38 assembly of the human genome and the effect allele was chosen to model BP increases. To be consistent with prior studies evaluating polygenic risks,^11^ the two polygenic risk scores were divided into 3 categories of genetic risk according to percentile cutoffs (≤20, 21-79, ≥80) labeled as low, intermediate and high PSH.

### Outcome ascertainment

The battery of cognitive tests administered by the UK Biobank was designed to be completed unsupervised. Although study staff members were present in the clinic during the test administration, participants were expected to work through the cognitive assessments independently. A detailed description of the cognitive tests administered by the UK Biobank is available elsewhere and summarized in Supplemental Table 2.^14^ Briefly, the cognitive assessments were designed to assess different cognitive domains in a short amount of time and to be completed without supervision on a touchscreen computer. At baseline, almost all participants completed the Pairs Memory test, a test of visual memory, and the Reaction Time test, a measure of processing speed. A portion of enrolled study participants also completed tests of working memory (Numeric Memory test), prospective memory (Prospective Memory), and verbal and numerical reasoning (Fluid Intelligence).

### Covariates

Age was calculated using the date of birth provided by study participants during the baseline visit. Sex and ancestry were determined using genetic information. Vascular risk factors and comorbidities were identified by combining baseline questionnaires, ICD 9/10 codes, and electronic health record data.

### Statistical analysis

We present discrete variables as counts (percentage [%]) and continuous variables as mean (standard deviation [SD]) or median (interquartile range [IQR]), as appropriate. All variables were evaluated to identify missing values and data entry errors. Unadjusted comparisons were made using chi-square tests for discrete variables and t or Wilcoxon Rank Sum tests, as appropriate, for continuous variables.

#### General cognitive ability score

Following prior reports,^15,16^ all five cognitive metrics were entered into a principal components analysis after appropriate transformation of non-normal distributed metrics (values of the Pairs Memory test were (log (x+1) transformed and values of the reaction time were log-transformed). The normalized values of the first unrotated principal component were saved and used as general cognitive ability scores, where higher values represent better cognitive ability (i.e. increased speed and/or accuracy).

#### Primary analysis

Our primary analysis was restricted to study participants of European ancestry, used the polygenic risk score built with variants related to systolic BP, and fitted multivariable linear regression to test for association between categories of PSH and the general cognitive ability score. The PSH categories were entered into the model as dummy-coded variables, generating one effect estimate for each of two possible comparisons (low versus intermediate and low versus high PSH). A single test for trend across these 2 comparisons was used to test the hypothesis that the association between PSH and general cognitive ability score changes in a linear fashion across categories of genetic risk. Following standard practices in statistical genetics,^17^ our primary modeling strategy included age, sex and genetic principal components 1 through 4 as covariates (Model 1). These covariates are included in regression models for precision, not to account for possible confounding. Because genetic risk variants are randomly distributed during meiosis, variant-disease associations are unlikely to be confounded by post-partum exposures.^18^

#### Sensitivity analyses

In sensitivity analyses, we evaluated other models: no covariates (Model 2); inclusion of age, sex, diabetes, hyperlipidemia, smoking and principal components 1 through 4 (Model 3); and variables included in Model 3 plus history of myocardial infarction (Model 4). Formal diagnosis of hypertension and observed BP were not included as covariates as they are likely to be mediators, rather than confounders, of the association of interest. All models used a complete data approach, where observations with missing data for any of the variables included in the model are dropped from the analysis.

#### Secondary analyses

In secondary analyses, we repeated the analyses described above in study participants of all race/ethnic groups and evaluated each cognitive test separately, fitting linear, or logistic regression models, as appropriate, and including the covariates described for Model 1 above.

#### Effect modification

We added product terms to our regression models to evaluate whether the association between PSH and cognitive performance was modified by age, sex or hypertension

#### Statistical significance and software

We declared statistical significance at p<0.05 (2-tailed) for the single statistical test, captured by our primary analysis, required to test the hypothesis that a higher PSH is associated with lower general cognitive ability score. Statistical analyses were completed in R 4.2.^19^ Genetic analyses were completed in PLINK 1.9.^20^

### Data availability statement

UK Biobank data are available through a procedure described at http://www.ukbiobank.ac.uk/using-the-resource/. Data were accessed using project application number 58743.

## RESULTS

Out of a total of 488,377 study participants enrolled in the UK Biobank with available genomic data, 48,118 (9.9%) completed all 5 cognitive tests administered during the baseline interview (Supplemental Table 3). Of these, 42,080 (96.2%) were of European ancestry (mean age 56.7 [SD 8.2], male sex 19,255 [45.8%]) and were included in primary analyses (Table 1). As expected, given the focus on middle-aged persons of the UK Biobank, the prevalence of vascular risk factors was relatively low, including hypertension 29.5% (n=12,396), hyperlipidemia 13.6% (n=5,708), diabetes 4.6% (n=1,920) and current smoking 9.5% (n=3,991). In addition, there were 1,594 study participants from other race/ethnic groups, including 332 (0.7%) who were Black, 976 (2%) who were Asian, and 286 (0.6%) who were of other race/ethnic groups (Supplemental Table 3). In terms of the general cognitive ability score, the first principal component of the principal component analysis capturing all 5 cognitive tests explained 38% of their combined variance, in line with prior findings.^15,16^

**Table 1.** Population characteristics for participants of European ancestry who completed all five cognitive tests at baseline.

| **Variable, n (%)** | **All**<br>**Categories of**<br>**genetic risk**<br>**n=42,080** | **Low**<br>**polygenic risk**<br>**for**<br>**hypertension\***<br>**n=8,417** | **Intermediate**<br>**polygenic risk**<br>**for**<br>**hypertension\***<br>**n=25,247** | **High**<br>**polygenic risk**<br>**for**<br>**hypertension\***<br>**n=8,416** | **P value**<br>**Comparison**<br>**across genetic**<br>**risk categories** |
| --- | --- | --- | --- | --- | --- |
| **Demographics** |  |  |  |  |  |
| **Age** | 56.7 (8.2) | 56.7 (8.3) | 56.7 (8.2) | 56.8 (8.2) | 0.62 |
| **Male sex** | 19,255 (45.8) | 3,894 (46.3) | 11,554 (45.8) | 3,775 (44.9) | 0.17 |
| **Vascular risk factors** |  |  |  |  |  |
| **Hypertension** | 12,396 (29.5) | 1,890 (22.5) | 7,323 (29.1) | 3,170 (37.7) | <0.001 |
| **Hyperlipidemia** | 57,08 (13.6) | 1,009 (12.0) | 3,444 (13.7) | 1,249 (14.9) | <0.001 |
| **Diabetes** | 1,920 (4.6) | 350 (4.2) | 1,135 (4.5) | 432 (5.1) | 0.008 |
| **Current smoker** | 3,991 (9.5) | 778 (9.3) | 2,413 (9.6) | 794 (9.5) | 0.81 |
| **Prior smoker** | 14,961 (35.7) | 2,962 (35.4) | 8,965 (35.7) | 3,005 (35.9) |  |
| **BMI, mean (SD)** | 27.37 (4.7) | 27.48 (4.7) | 27.35 (4.7) | 27.31 (4.6) | 0.038 |
| **Comorbidities** |  |  |  |  |  |
| **Myocardial infarction** | 1255 (3.0) | 194 (2.3) | 769 (3.1) | 289 (3.4) | <0.001 |
| **Atrial fibrillation** | 1808 (4.3) | 334 (4.0) | 1069 (4.2) | 401 (4.8) | 0.031 |
| **Admission BP, mean (SD)** |  |  |  |  |  |
| **Systolic BP** | 139.3 (18.8) | 134.9 (17.9) | 139.2 (18.6) | 143.7 (19.4) | <0.001 |
| **Diastolic BP** | 82.53 (10.01) | 80.80 (9.79) | 82.53 (9.94) | 84.27 (10.12) | <0.001 |

Abbreviations: BMI = body mass index; SD = standard deviation; BP = blood pressure.
\* Polygenic risk for hypertension estimated using genetic risk variants for systolic blood pressure.

### Polygenic susceptibility to hypertension and observed blood pressure and hypertension

Higher PSH, modeled through a polygenic risk score containing genetic risk variants for systolic BP, was significantly associated with higher observed BP as well as higher prevalence of formally diagnosed hypertension (Table 1). In study participants with low, intermediate and high PSH, the mean systolic BP was 134.9 (SD 17.9), 139.2 (18.6) and 143.7 (SD 19.4), respectively (unadjusted p<0.001). Similarly, in study participants with low, intermediate and high PSH, the mean diastolic BP was 80.8 (SD 9.8), 82.5 (SD 9.9) and 84.3 (SD 10.1), respectively (unadjusted p<0.001). The prevalence of formally diagnosed hypertension followed a similar pattern, being present in 22.5% (n=1,890), 29.1% (n=7,323) and 37.7% (n=3,170) of study participants with low, intermediate and high PSH, respectively (unadjusted p<0.001). Similar associations were found when evaluating study participants of all race/ethnic groups (Supplemental Table 3) and when modeling PSH through a polygenic risk score containing genetic risk variants for diastolic BP (data not shown).

### Polygenic susceptibility to hypertension and general cognitive ability

We found that a higher PSH was associated with a lower (worse) general cognitive ability score in this cohort (Tables 2 and 3). Our primary analysis focused on Europeans and used a polygenic risk score calculated using genetic risk variants for systolic BP. In this setting, the mean general cognitive ability score (represented by the normalized first principal component of all 5 cognitive tests) in study participants with low, intermediate and high PSH was 0.08 (SD 0.96), 0.04 (SD 0.97) and 0.01 (SD 0.96), respectively (unadjusted p<0.001, Table 2). Similar significant associations were found in multivariable linear regression models adjusting for age, sex and principal components (Table 3, Model 1): compared to study participants with low PSH, those with intermediate and high PSH had reductions of 3.4% (beta -0.034, SE 0.012) and 6.6% (beta - 0.066, SE 0.014) in the general cognitive ability score (test for trend p<0.001). These results remained significant when not including covariates (Model 2, test for trend p<0.001), when including hyperlipidemia, diabetes and smoking (Model 3, test for trend p<0.001) and when adding history of myocardial infarction to Model 3 (Model 4, test for trend p<0.006). These results also remained significant in secondary analyses that included study participants of all race/ethnic groups (Supplemental Table 4) and that used genetic risk variants for diastolic BP to model PSH (Table 2). Interaction analyses did not reveal any significant effect modification by age, sex or hypertension status (for all tests, interaction p>0.05, data not shown).

**Table 2.**
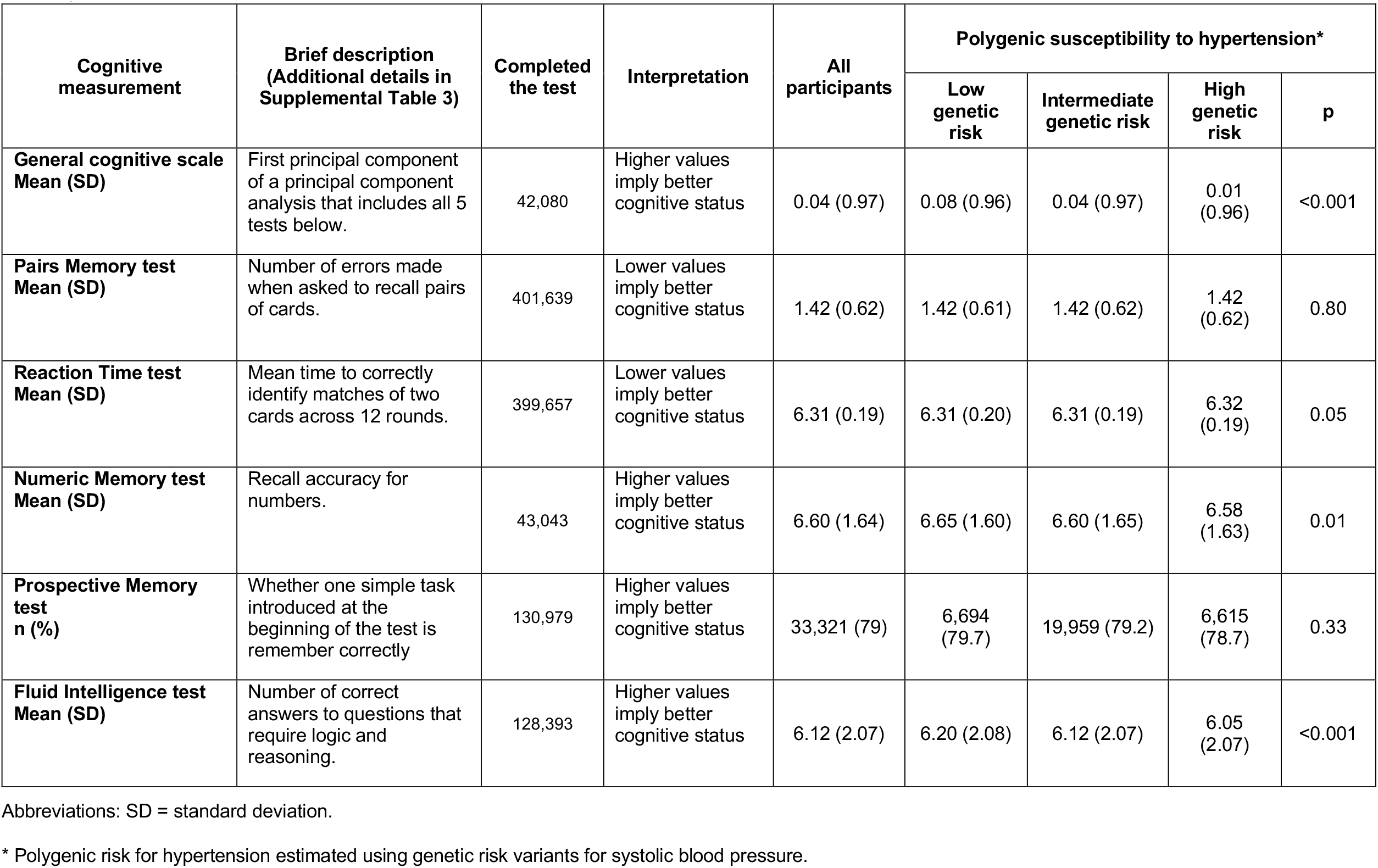
Cognitive measurements: descriptive statistics and unadjusted comparisons across categories of genetic risk in participants of European ancestry.

| Cognitive measurement | Brief description<br>(Additional details in Supplemental Table 3) | Completed the test | Interpretation | All participants | Polygenic susceptibility to hypertension* |  |  |  |
| --- | --- | --- | --- | --- | --- | --- | --- | --- |
|  |  |  |  |  | Low genetic risk | Intermediate genetic risk | High genetic risk | p |
| **General cognitive scale Mean (SD)** | First principal component of a principal component analysis that includes all 5 tests below. | 42,080 | Higher values imply better cognitive status | 0.04 (0.97) | 0.08 (0.96) | 0.04 (0.97) | 0.01 (0.96) | <0.001 |
| **Pairs Memory test Mean (SD)** | Number of errors made when asked to recall pairs of cards. | 401,639 | Lower values imply better cognitive status | 1.42 (0.62) | 1.42 (0.61) | 1.42 (0.62) | 1.42 (0.62) | 0.80 |
| **Reaction Time test Mean (SD)** | Mean time to correctly identify matches of two cards across 12 rounds. | 399,657 | Lower values imply better cognitive status | 6.31 (0.19) | 6.31 (0.20) | 6.31 (0.19) | 6.32 (0.19) | 0.05 |
| **Numeric Memory test Mean (SD)** | Recall accuracy for numbers. | 43,043 | Higher values imply better cognitive status | 6.60 (1.64) | 6.65 (1.60) | 6.60 (1.65) | 6.58 (1.63) | 0.01 |
| **Prospective Memory test n (%)** | Whether one simple task introduced at the beginning of the test is remember correctly | 130,979 | Higher values imply better cognitive status | 33,321 (79) | 6,694 (79.7) | 19,959 (79.2) | 6,615 (78.7) | 0.33 |
| **Fluid Intelligence test Mean (SD)** | Number of correct answers to questions that require logic and reasoning. | 128,393 | Higher values imply better cognitive status | 6.12 (2.07) | 6.20 (2.08) | 6.12 (2.07) | 6.05 (2.07) | <0.001 |

Abbreviations: SD = standard deviation.
\* Polygenic risk for hypertension estimated using genetic risk variants for systolic blood pressure.

**Table 3.**
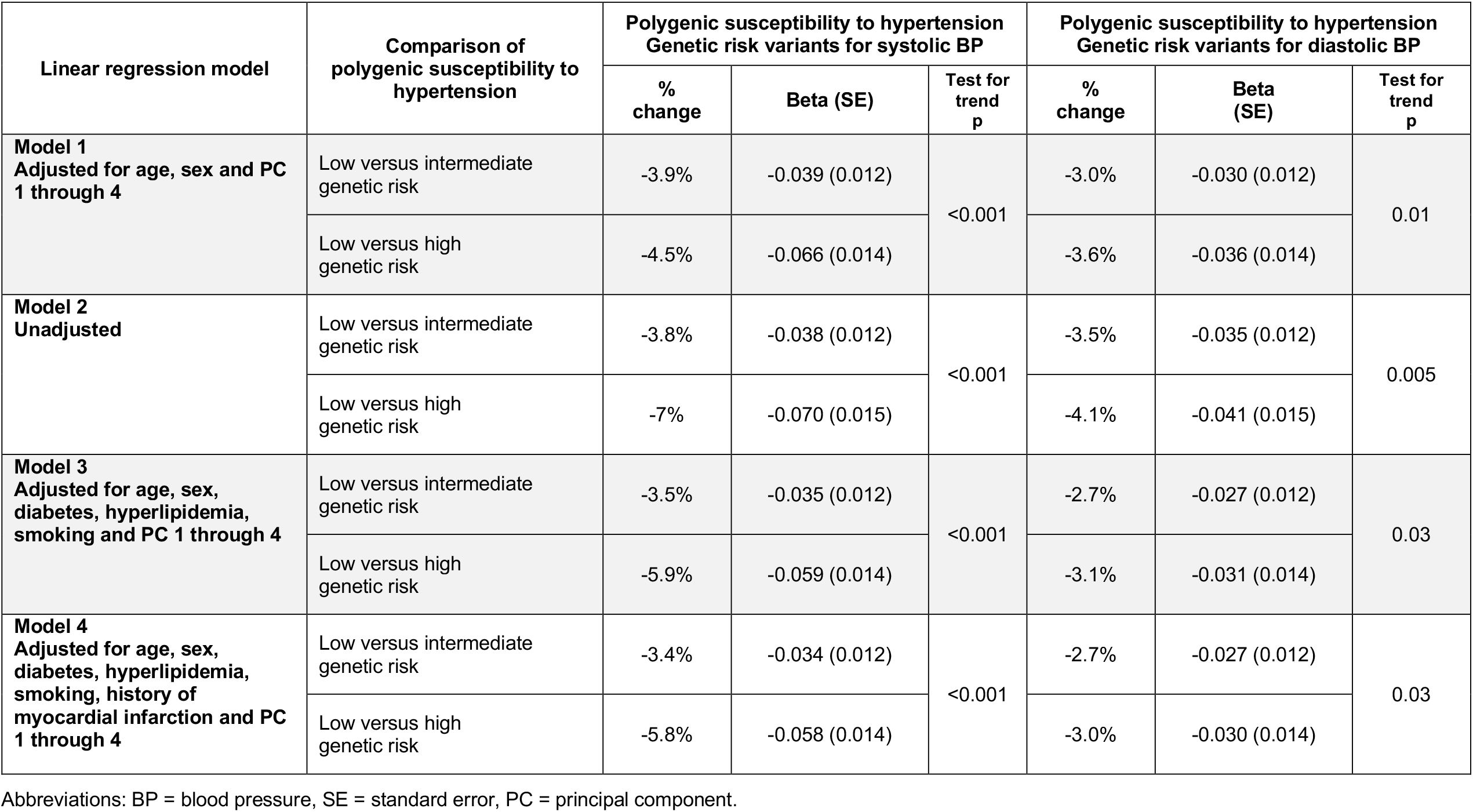
Association between polygenic susceptibility to hypertension and baseline cognitive status in participants of European ancestry.

### Polygenic susceptibility to hypertension and performance in each cognitive test

When separately evaluating each of the five cognitive tests captured by the general cognitive ability score, we found that the relationship between PSH and cognitive performance was driven by specific tests (Table 4). Our primary analysis focused on Europeans, used a polygenic risk score calculated using genetic risk variants for systolic BP, and fitted multivariable regression analyses adjusting for age, sex and principal components 1 through 4. We found that a higher PSH was associated with worse performance in the following tests (Table 4): Reaction Time (low versus high genetic risk, beta 0.033, SE 0.015, test for trend p=0.02), Numeric Memory (low versus high genetic risk, beta -0.038, SE 0.014, test for trend p=0.006) and Fluid Intelligence (low versus high genetic risk, beta -0.069, SE 0.011, test for trend p<0.001). No associations were found for the Pairs Memory and Prospective Memory tests (both instances, test for trend p>0.05). Consistent results were found in secondary analyses including study participants of all race/ethnic groups (Supplemental Table 5), although the results for Prospective Memory were in the same direction and magnitude but statistically non-significant (p=0.11). Of note, the results were less consistent in secondary analyses that used genetic risk variants for diastolic BP to model PSH, only yielding a significant association for Reaction Time (p=0.03, Table 4). Interaction analyses did not reveal any significant effect modification by age, sex or hypertension status.

**Table 4.**
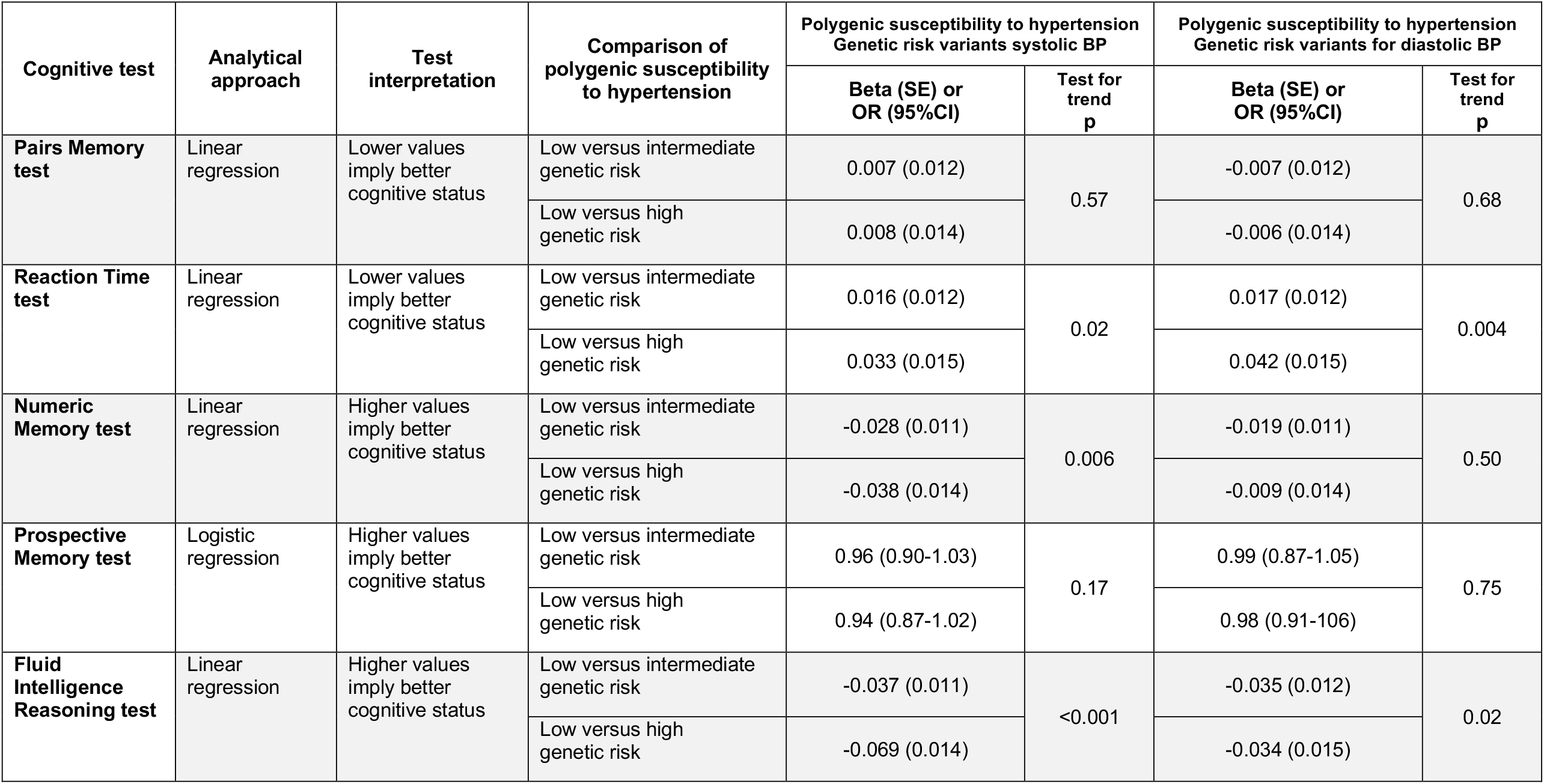
Association between polygenic susceptibility to hypertension and results of 5 different cognitive tests in participants of European ancestry.

| Cognitive test | Analytical approach | Test interpretation | Comparison of polygenic susceptibility to hypertension | Polygenic susceptibility to hypertension<br>Genetic risk variants systolic BP |  | Polygenic susceptibility to hypertension<br>Genetic risk variants for diastolic BP |  |
| --- | --- | --- | --- | --- | --- | --- | --- |
|  |  |  |  | Beta (SE) or OR (95%CI) | Test for trend p | Beta (SE) or OR (95%CI) | Test for trend p |
| **Pairs Memory test** | Linear regression | Lower values imply better cognitive status | Low versus intermediate genetic risk | 0.007 (0.012) | 0.57 | -0.007 (0.012) | 0.68 |
|  |  |  | Low versus high genetic risk | 0.008 (0.014) |  | -0.006 (0.014) |  |
| **Reaction Time test** | Linear regression | Lower values imply better cognitive status | Low versus intermediate genetic risk | 0.016 (0.012) | 0.02 | 0.017 (0.012) | 0.004 |
|  |  |  | Low versus high genetic risk | 0.033 (0.015) |  | 0.042 (0.015) |  |
| **Numeric Memory test** | Linear regression | Higher values imply better cognitive status | Low versus intermediate genetic risk | -0.028 (0.011) | 0.006 | -0.019 (0.011) | 0.50 |
|  |  |  | Low versus high genetic risk | -0.038 (0.014) |  | -0.009 (0.014) |  |
| **Prospective Memory test** | Logistic regression | Higher values imply better cognitive status | Low versus intermediate genetic risk | 0.96 (0.90-1.03) | 0.17 | 0.99 (0.87-1.05) | 0.75 |
|  |  |  | Low versus high genetic risk | 0.94 (0.87-1.02) |  | 0.98 (0.91-1.06) |  |
| **Fluid Intelligence Reasoning test** | Linear regression | Higher values imply better cognitive status | Low versus intermediate genetic risk | -0.037 (0.011) | <0.001 | -0.035 (0.012) | 0.02 |
|  |  |  | Low versus high genetic risk | -0.069 (0.014) |  | -0.034 (0.015) |  |

## DISCUSSION

We report the results of a genetic study that evaluated whether a higher polygenic susceptibility to hypertension is associated with cognitive performance in community-dwelling, middle-aged adults without dementia or stroke enrolled in a large population study. We found that a higher PSH, modeled via polygenic risk scores, was significantly associated with worse cognitive performance in a battery of five tests that, while simple in their concept and execution, have been shown to correlate well with traditional, more detailed cognitive tests.^15^ These associations were consistent when modeling PSH using genetic risk variants related to both systolic and diastolic BP and were robust to different modeling strategies, retaining their significance when evaluating participants of any race/ethnic group, when adjusting for multiple vascular risk factors and history of cardiovascular disease. We did not find evidence for synergistic effects between PSH and other known risk factors for cognitive decline and dementia.

Our results extend the existing body of evidence indicating that hypertension is an important treatable risk factor for cognitive decline and dementia. Coupled with its well-established role as a risk factor for stroke, the links with cognitive decline and dementia makes hypertension an appealing treatment target to preserve brain health.^21^ The enthusiasm for this strategy is bolstered by the extensive experience with, and numerous drugs for, the treatment of blood pressure in different clinical contexts, that run the gamut from primary prevention to secondary prevention, to treatment of acute cardiovascular events.^22^ As mentioned before, the SPRINT MIND clinical trial found that a strict systolic BP target <120 mmHg reduced the risk of cognitive decline as well as the composite risk of probable dementia and cognitive decline.^3^ Moreover, an ancillary neuroimaging study to SPRINT MIND found that this target BP goal was also associated with a slower progression of white matter hyperintensities,^4^ a well-established marker of silent cerebrovascular disease measured in brain magnetic resonance imaging.^5,6^ In light of these important clinical and neuroimaging findings, and given the prominent role of genetic variation in determining the level and response to treatment of BP, genetic analyses constitute a unique tool to advance our understanding of the role of hypertension in cognition.^23^

The results of the present study provide important new knowledge about the role of PSH in brain health by highlighting the crucial role played by this genetic predisposition in the development of mild cognitive changes. Prior research showed that a higher PSH is associated with a higher risk of dementia of any cause in the UK Biobank population.^13^ As expected, the total number of cases and overall risk of dementia in this prior study was low due to the relatively young age of this cohort. Our results provide an important piece of new evidence by showing that PSH influences cognitive performance in persons who have not been diagnosed with dementia. One important implication of these findings is extending the importance of genetically-determined hypertension to a much larger population, as subtle cognitive changes can have important functional implications even in non-demented persons.^24^ In addition, because subtle cognitive changes constitute a risk factor for dementia, our results emphasize the importance of addressing BP control as early as possible during middle age.

Our findings also address the unmet need to develop novel precision medicine strategies for the ultra-early identification of middle age persons at high risk of developing unfavorable BP profiles that ultimately contribute to poor brain health. Despite the existing evidence supporting the importance of early and sustained BP control, the optimization of cardiovascular health during middle age remains a challenge due to the lack of regular interactions with the health care system during this period. Because genetic information is available since birth and remains mostly unchanged throughout life,^18^ polygenic risk data allows the identification of high-risk persons long before a formal diagnosis of hypertension is made in the clinic.^25^ The implementation of such ultra-early risk stratification strategies would allow the development of interventions aimed at delaying, or altogether preventing, hypertension, rather than its downstream clinical consequences (like cognitive decline, dementia and stroke). Given the increasingly prominent role played by the social determinants of health in brain and cardiovascular health,^26^ these omic-based approaches to precision medicine could be amplified by combining genomic with socioeconomic and cultural data.^27^

A number of recent scientific developments will significantly facilitate the design and implementation of the genomic-based precision medicine strategies outlined above. First, clinical trials are increasingly including genomic stratification as part of their scientific goals. As an example, pre-specified genetic studies of the *Further Cardiovascular Outcomes Research With PCSK9 Inhibition in Subjects With Elevated Risk* (FOURIER) and *Evaluation of Cardiovascular Outcomes After an Acute Coronary Syndrome During Treatment With Alirocumab* (ODYSSEY OUTCOMES) trials demonstrated that persons with high genetic risk of cardiovascular disease obtained the greatest benefit from treatment with protein convertase subtilisin/kexin type 9 (PCSK9) inhibitors.^28,29^ Importantly, in these analyses the combination of genetic risk and clinical predictive scores appeared better than any of these separately.^43^ Second, the recent introduction of large-scale precision medicine studies, that include genomic information, will significantly accelerate research on this front. Developed and maintained by the National Institutes of Health, the *All of Us* Research Program plans to enroll a diverse group of at least 1 million Americans to propel biomedical research and reduce health disparities.^30^ In addition to making all generated data publicly available for research purposes, a central goal of *All of Us* is to return genome-wide data to enrolled study participants. Third, the decreasing cost of genotyping and the rapidly growing number of persons genotyped by direct-to-consumer companies (currently in the several millions worldwide) will make the adoption of these strategies much easier compared to a decade ago.^31^

The most important strengths of our study are its large sample size and the focus on middle age persons who are generally healthy, a population group likely to benefit from genomic-based precision medicine strategies. As for limitations, first, it should be noted that the cognitive tests administered by the UK Biobank are less comprehensive than other, more traditional batteries regularly used in the clinic or in cognitive research. As mentioned before, the simplicity of these tests stems from the need to balance depth and precision with the mandate to acquire as much data about human disease as possible in this large-scale, general-purpose population study. Importantly, prior research in patients who both participated in the UK Biobank and underwent extensive cognitive testing shows that the general cognitive ability score used here correlates well or very well with standard cognitive tests used in the clinic.^15^ Another important limitation of our study is the extreme predominance of participants of European ancestry, which limits the portability of our findings to other race/ethnic groups. It is therefore imperative to externally validate these results in non-white populations, to reduce the large underrepresentation of minority populations in genetic studies.

In conclusion, we report the results of a genetic study that evaluated whether a higher polygenic susceptibility to hypertension is associated with cognitive performance in community-dwelling, middle-aged adults without dementia or stroke enrolled in a large population study. We found that a higher PSH, modeled via polygenic risk scores, was significantly associated with worse cognitive performance in a battery of five simple tests designed to be administered in the setting of a large population study. These findings extend the existing evidence on the important role of PSH in cognition by showing that this genetic predisposition can influence cognitive performance in persons without dementia. In addition, our results provide important support to follow-up research focused on evaluating whether genomic information can help identify middle age persons at high risk of developing hypertension and adverse brain health profiles.

## Data Availability

All data produced in the present study are available upon reasonable request to the authors.

https://www.ukbiobank.ac.uk/

## Acknowledgments

None.

