## Supplemental Data for "Polygenic Susceptibility to Hypertension is Associated with Worse Cognitive Performance in Middle-Aged Persons without Dementia"

Julian N. Acosta MD, Cameron P. Both BS, Richa Sharma MD,

Seyedmehdi Payabvash MD, Adam De Havenon MD, Kevin N. Sheth MD,

Thomas M. Gill MD, Guido J. Falcone MD ScD MPH

**Supplemental Table 1. Single nucleotide polymorphisms included in the polygenic risk score.**

| CHR | BP | SNP | A1 | A2 | Beta_SBP | se_SBP | Beta_DBP | se_DBP |
|---|---|---|---|---|---|---|---|---|
| 1 | 1687482 | rs2076328 | T | G | -0.2824 | 0.0498 | -0.0965 | 0.0292 |
| 1 | 2187085 | rs260508 | T | G | 0.1297 | 0.0441 | 0.0854 | 0.0323 |
| 1 | 3328659 | rs2493292 | T | C | 0.2686 | 0.068 | 0.1674 | 0.0401 |
| 1 | 6278414 | rs709209 | A | G | 0.0935 | 0.0528 | -0.0271 | 0.0308 |
| 1 | 7739250 | rs4908678 | T | C | -0.1203 | 0.0484 | -0.1065 | 0.0285 |
| 1 | 8422676 | rs2252865 | T | C | 0.159 | 0.0434 | 0.0976 | 0.0318 |
| 1 | 9441949 | rs9662255 | A | C | -0.1883 | 0.0482 | -0.0156 | 0.0283 |
| 1 | 10796866 | rs880315 | T | C | -0.5218 | 0.0499 | -0.2524 | 0.0293 |
| 1 | 11862778 | rs17367504 | A | G | 0.7774 | 0.0639 | 0.4715 | 0.0377 |
| 1 | 15798197 | rs3820068 | A | G | 0.3361 | 0.0596 | 0.1279 | 0.0349 |
| 1 | 22577371 | rs2807337 | T | C | 0.1551 | 0.0423 | 0.0587 | 0.031 |
| 1 | 23442265 | rs150266910 | T | C | 0.0726 | 0.0549 | -0.0572 | 0.0403 |
| 1 | 25030470 | rs6686889 | T | C | 0.0854 | 0.0531 | 0.115 | 0.0312 |
| 1 | 27284913 | rs79598313 | T | C | 0.456 | 0.1367 | 0.1923 | 0.0994 |
| 1 | 28734372 | rs143167197 | A | G | -0.3388 | 0.0834 | -0.0493 | 0.0608 |
| 1 | 29549216 | rs1565716 | A | G | 0.3142 | 0.0837 | 0.16 | 0.0614 |
| 1 | 38298207 | rs9729719 | A | G | 0.1015 | 0.053 | -0.0344 | 0.0311 |
| 1 | 41865293 | rs11210029 | A | G | -0.1608 | 0.0476 | -0.0934 | 0.0281 |
| 1 | 42408070 | rs7515635 | T | C | 0.2382 | 0.0463 | 0.109 | 0.0273 |
| 1 | 43037556 | rs72659998 | T | C | -0.1037 | 0.0569 | 0.0535 | 0.0417 |
| 1 | 43856410 | rs839755 | A | C | -0.1499 | 0.0434 | -0.0791 | 0.0318 |
| 1 | 46027355 | rs512083 | T | C | -0.0998 | 0.0419 | 0.0453 | 0.0307 |
| 1 | 46541679 | rs12142296 | T | G | -0.2038 | 0.0633 | -0.1959 | 0.0463 |
| 1 | 48109225 | rs4926923 | T | C | 0.1997 | 0.0834 | 0.1459 | 0.0492 |
| 1 | 49052423 | rs11579440 | T | C | 0.2794 | 0.0653 | 0.1546 | 0.0383 |
| 1 | 51021867 | rs147696085 | A | G | -0.0739 | 0.0797 | 0.1416 | 0.047 |
| 1 | 51527684 | rs6681713 | T | G | 0.2032 | 0.163 | 0.3834 | 0.1194 |
| 1 | 56576924 | rs112557609 | A | G | 0.2383 | 0.0489 | 0.083 | 0.0287 |
| 1 | 57008778 | rs2404715 | T | C | -0.1997 | 0.0797 | 0.0016 | 0.0469 |
| 1 | 59663341 | rs60199046 | A | G | 0.1782 | 0.0507 | -0.0758 | 0.0297 |
| 1 | 67071356 | rs20354 | T | G | 0.1836 | 0.0612 | 0.0285 | 0.0449 |
| 1 | 78450517 | rs34517439 | A | C | 0.0172 | 0.0736 | -0.165 | 0.0537 |
| 1 | 86040107 | rs12034319 | A | G | 0.1072 | 0.0514 | 0.0085 | 0.0376 |
| 1 | 86822231 | rs385437 | A | G | 0.168 | 0.0581 | 0.0314 | 0.0424 |
| 1 | 88651771 | rs10923038 | A | C | 0.1279 | 0.0481 | 0.062 | 0.0283 |
| 1 | 89360158 | rs10922502 | A | G | -0.2283 | 0.0483 | -0.105 | 0.0284 |
| 1 | 90228519 | rs2065152 | T | C | 0.1855 | 0.0478 | 0.1374 | 0.0281 |
| 1 | 94051350 | rs7514579 | A | C | 0.1443 | 0.05 | 0.04 | 0.0367 |
| 1 | 94730954 | rs17396055 | A | G | -0.1654 | 0.0462 | -0.07 | 0.0338 |
| 1 | 113190807 | rs17030613 | A | C | -0.3923 | 0.056 | -0.2359 | 0.033 |
| 1 | 113216543 | rs2932538 | A | G | -0.3435 | 0.0527 | -0.237 | 0.0311 |
| 1 | 154245917 | rs13796 | T | C | -0.0984 | 0.0705 | -0.1446 | 0.0413 |
| 1 | 156129796 | rs76719272 | T | C | -0.1708 | 0.0651 | -0.0415 | 0.0476 |
| 1 | 164740099 | rs2171690 | T | C | 0.151 | 0.0461 | 0.1371 | 0.0272 |
| 1 | 167367193 | rs7524019 | T | C | 0.1516 | 0.0466 | 0.0729 | 0.0275 |
| 1 | 169201567 | rs2157597 | T | C | -0.0856 | 0.049 | 0.0034 | 0.0288 |
| 1 | 172357441 | rs12405515 | T | G | -0.1713 | 0.0461 | -0.1617 | 0.0272 |
| 1 | 176634724 | rs12118102 | A | G | 0.2941 | 0.0939 | 0.0111 | 0.0687 |
| 1 | 179571862 | rs150816167 | T | C | -0.0442 | 0.1247 | -0.2378 | 0.0727 |
| 1 | 180131640 | rs10913934 | T | G | 0.1302 | 0.0439 | -0.0255 | 0.0322 |
| 1 | 180859368 | rs1043069 | T | G | 0.1287 | 0.0446 | 0.0445 | 0.0327 |
| 1 | 183058452 | rs41475048 | A | G | -0.1144 | 0.0549 | -0.0869 | 0.0323 |
| 1 | 184585182 | rs4651224 | T | C | 0.186 | 0.0433 | 0.1277 | 0.0317 |
| 1 | 197297417 | rs12042924 | T | C | -0.1235 | 0.0427 | -0.0202 | 0.0313 |
| 1 | 201735913 | rs882624 | T | C | 0.0116 | 0.0458 | -0.1423 | 0.0335 |
| 1 | 203109801 | rs33996239 | T | C | -0.3263 | 0.1012 | -0.1663 | 0.074 |
| 1 | 204518842 | rs4245739 | A | C | 0.1702 | 0.0525 | 0.1364 | 0.0309 |
| 1 | 207220800 | rs2629665 | A | C | -0.205 | 0.0476 | -0.1199 | 0.028 |
| 1 | 207919748 | rs2761436 | T | C | 0.1927 | 0.0461 | 0.0311 | 0.0271 |
| 1 | 217718789 | rs12408022 | T | C | 0.2511 | 0.0532 | 0.1486 | 0.0313 |
| 1 | 219753509 | rs2820443 | T | C | 0.2272 | 0.0457 | 0.0317 | 0.0335 |
| 1 | 221358796 | rs9431431 | A | G | -0.1703 | 0.0506 | -0.1088 | 0.0298 |
| 1 | 227250775 | rs73091767 | T | C | -0.0906 | 0.0516 | -0.0874 | 0.0304 |
| 1 | 230848702 | rs2004776 | T | C | 0.3368 | 0.0542 | 0.2356 | 0.0319 |
| 1 | 243472801 | rs6429422 | T | G | -0.1434 | 0.0492 | -0.1919 | 0.029 |

| | | | | | | | | |
|---|---|---|---|---|---|---|---|---|
| 2 | 9298590 | rs2175337 | A | C | 0.1118 | 0.0432 | -0.0678 | 0.0316 |
| 2 | 18975439 | rs67720684 | A | C | 0.2242 | 0.0512 | 0.0676 | 0.0375 |
| 2 | 19730845 | rs1344653 | A | G | -0.1568 | 0.0456 | 0.068 | 0.0268 |
| 2 | 20878820 | rs7255 | T | C | -0.0707 | 0.0469 | 0.0752 | 0.0276 |
| 2 | 21423532 | rs66774912 | A | G | -0.1582 | 0.0674 | 0.0366 | 0.0398 |
| 2 | 23950200 | rs10779936 | A | G | -0.1093 | 0.0505 | 0.0174 | 0.0298 |
| 2 | 25139596 | rs55701159 | T | G | 0.2999 | 0.0742 | 0.1466 | 0.0435 |
| 2 | 26914364 | rs1275988 | T | C | -0.5157 | 0.0466 | -0.2792 | 0.0274 |
| 2 | 27887034 | rs9678851 | A | C | -0.1135 | 0.0474 | -0.0585 | 0.0278 |
| 2 | 28635740 | rs7562 | T | C | 0.1555 | 0.047 | 0.0832 | 0.0276 |
| 2 | 34679626 | rs1607644 | A | G | -0.1269 | 0.0427 | -0.1343 | 0.0313 |
| 2 | 37517566 | rs13420463 | A | G | 0.2751 | 0.0555 | 0.1273 | 0.0326 |
| 2 | 40567743 | rs4952611 | T | C | -0.2135 | 0.0487 | -0.1342 | 0.0286 |
| 2 | 42352567 | rs11681462 | A | C | -0.1806 | 0.0583 | -0.1257 | 0.0343 |
| 2 | 43167878 | rs76326501 | A | C | 0.5511 | 0.0829 | 0.36 | 0.0487 |
| 2 | 43716933 | rs35590893 | A | G | -0.1706 | 0.0477 | -0.0595 | 0.0349 |
| 2 | 46363336 | rs11690961 | A | C | 0.0228 | 0.0722 | -0.1655 | 0.0424 |
| 2 | 50429861 | rs6545155 | T | C | 0.1442 | 0.0503 | 0.07 | 0.037 |
| 2 | 53025757 | rs10189186 | A | G | 0.1752 | 0.0459 | 0.0753 | 0.027 |
| 2 | 55279681 | rs2920899 | T | G | 0.1579 | 0.0529 | 0.0677 | 0.0386 |
| 2 | 55809054 | rs1975487 | A | G | -0.2206 | 0.0472 | -0.1737 | 0.0277 |
| 2 | 59315828 | rs6730325 | A | G | -0.1258 | 0.0428 | 3.00E-04 | 0.0314 |
| 2 | 61836235 | rs7608483 | A | C | 0.1652 | 0.047 | 0.1016 | 0.0277 |
| 2 | 64217786 | rs13014371 | T | C | -0.1602 | 0.0464 | -0.1086 | 0.0273 |
| 2 | 66104881 | rs2631669 | T | C | -0.0029 | 0.0416 | -0.0904 | 0.0305 |
| 2 | 66782467 | rs2300481 | T | C | 0.1887 | 0.0427 | 0.075 | 0.0313 |
| 2 | 68503044 | rs6731373 | A | G | 0.1785 | 0.0461 | 0.007 | 0.0338 |
| 2 | 69065841 | rs12052761 | A | G | -0.1301 | 0.0475 | -0.1232 | 0.0279 |
| 2 | 71627539 | rs3771371 | T | C | -0.1814 | 0.0464 | -0.0684 | 0.0273 |
| 2 | 72483329 | rs10193543 | T | G | 0.2859 | 0.0629 | 0.1996 | 0.0376 |
| 2 | 73114352 | rs1876487 | A | C | -0.0825 | 0.0545 | -0.1105 | 0.0319 |
| 2 | 85491365 | rs11689667 | T | C | 0.1189 | 0.0465 | -0.0348 | 0.0274 |
| 2 | 86326717 | rs72847885 | A | G | 0.1717 | 0.0452 | 0.0746 | 0.0331 |
| 2 | 96675166 | rs2579519 | T | C | -0.0054 | 0.0484 | -0.1499 | 0.0284 |
| 2 | 98357163 | rs4851462 | T | C | -0.0051 | 0.0437 | -0.0948 | 0.032 |
| 2 | 112769721 | rs28377357 | A | G | -0.1335 | 0.0454 | -0.0354 | 0.0332 |
| 2 | 114082175 | rs62158170 | A | G | 0.2939 | 0.0563 | 0.1836 | 0.0332 |
| 2 | 121440218 | rs10864859 | T | G | 0.3737 | 0.0886 | 0.286 | 0.0521 |
| 2 | 122000745 | rs6723509 | T | C | 0.2018 | 0.0588 | 0.1046 | 0.0432 |
| 2 | 127183454 | rs13001283 | A | G | 0.1232 | 0.0564 | 0.1484 | 0.0414 |
| 2 | 135632981 | rs4954192 | T | C | -0.1862 | 0.0481 | -0.1111 | 0.0284 |
| 2 | 138421227 | rs72844590 | T | G | 0.1893 | 0.0627 | 0.0179 | 0.0457 |
| 2 | 144146311 | rs7606205 | A | C | -0.1378 | 0.046 | -0.0994 | 0.0337 |
| 2 | 145646072 | rs1438896 | T | C | 0.2275 | 0.05 | 0.1912 | 0.0294 |
| 2 | 146272860 | rs34570306 | T | C | -0.1363 | 0.0433 | -0.0851 | 0.0317 |
| 2 | 146950908 | rs62169544 | A | G | -0.1318 | 0.0466 | -0.1101 | 0.0274 |
| 2 | 148572160 | rs12990959 | T | C | -0.1368 | 0.0498 | -0.1197 | 0.0293 |
| 2 | 152978341 | rs4664080 | A | G | -0.0883 | 0.0436 | 0.0334 | 0.032 |
| 2 | 153618773 | rs3175 | A | G | 0.1549 | 0.0501 | 0.0465 | 0.0295 |
| 2 | 161368213 | rs79523138 | A | G | -0.301 | 0.0731 | -0.0544 | 0.0535 |
| 2 | 162278233 | rs55732192 | T | G | -0.2807 | 0.0798 | -0.1224 | 0.047 |
| 2 | 164963486 | rs1446468 | T | C | -0.487 | 0.0468 | -0.2625 | 0.0275 |
| 2 | 165557318 | rs6712203 | T | C | -0.1943 | 0.0477 | -0.1137 | 0.0281 |
| 2 | 166250129 | rs2390258 | A | G | -0.1137 | 0.0448 | -0.1132 | 0.0329 |
| 2 | 169763148 | rs560887 | T | C | -0.078 | 0.0451 | 0.0362 | 0.0331 |
| 2 | 172381487 | rs151054210 | A | G | 0.073 | 0.0544 | -0.0809 | 0.0399 |
| 2 | 173965056 | rs6758859 | T | C | 0.0481 | 0.0477 | 0.098 | 0.028 |
| 2 | 174949358 | rs11694601 | A | G | -0.1422 | 0.047 | -0.0818 | 0.0277 |
| 2 | 179786068 | rs79146658 | T | C | -0.0045 | 0.086 | -0.2819 | 0.0505 |
| 2 | 180739450 | rs1486236 | A | C | -0.1058 | 0.0445 | 0.0044 | 0.0326 |
| 2 | 183224127 | rs16823124 | A | G | 0.2319 | 0.0497 | 0.1804 | 0.0292 |
| 2 | 187816321 | rs28558491 | T | C | -0.1419 | 0.0473 | -0.055 | 0.0347 |
| 2 | 189643316 | rs11901929 | A | G | 0.0938 | 0.0449 | -0.0057 | 0.0329 |
| 2 | 191439591 | rs7592578 | T | G | -0.3414 | 0.0603 | -0.1875 | 0.0354 |
| 2 | 201102905 | rs296797 | T | C | 0.2067 | 0.0467 | 0.0629 | 0.0275 |
| 2 | 204125426 | rs1469760 | T | C | -0.1282 | 0.0425 | -0.0065 | 0.0311 |
| 2 | 205077128 | rs2162003 | T | C | 0.1652 | 0.0507 | 0.1002 | 0.0297 |

| | | | | | | | | |
|---|---|---|---|---|---|---|---|---|
| 2 | 207996447 | rs1263671 | T | C | -0.1405 | 0.0645 | -0.1586 | 0.038 |
| 2 | 208526140 | rs55780018 | T | C | -0.3278 | 0.0488 | -0.1436 | 0.0286 |
| 2 | 211540507 | rs1047891 | A | C | -0.1647 | 0.0511 | -0.1028 | 0.0301 |
| 2 | 213188795 | rs12694277 | T | C | -0.1284 | 0.0462 | -0.0684 | 0.0338 |
| 2 | 217659266 | rs4674114 | A | G | -0.1366 | 0.0524 | -0.0322 | 0.0386 |
| 2 | 218668732 | rs1063281 | T | C | -0.2392 | 0.0483 | -0.1378 | 0.0283 |
| 2 | 219651349 | rs1996992 | T | G | -0.1292 | 0.0957 | -0.2953 | 0.0703 |
| 2 | 220362557 | rs12474050 | T | C | 0.0504 | 0.0486 | 0.1025 | 0.0286 |
| 2 | 227100698 | rs2972146 | T | G | 0.2486 | 0.0476 | 0.1395 | 0.0281 |
| 2 | 230629138 | rs1044822 | T | C | -0.1852 | 0.0608 | -0.0921 | 0.0446 |
| 2 | 238227594 | rs12052878 | A | G | -0.0697 | 0.0442 | 0.0231 | 0.0324 |
| 2 | 239864732 | rs4507125 | A | C | -0.1046 | 0.0554 | -0.1418 | 0.0326 |
| 2 | 242344695 | rs139354822 | T | C | 0.442 | 0.1248 | 0.3155 | 0.0906 |
| 3 | 7489993 | rs9865843 | A | G | -0.1161 | 0.0419 | -0.0856 | 0.0307 |
| 3 | 11290122 | rs347591 | T | G | 0.2842 | 0.0489 | 0.1472 | 0.0288 |
| 3 | 13826854 | rs729639 | T | C | -0.0663 | 0.0442 | 0.0498 | 0.0324 |
| 3 | 14958126 | rs11128722 | A | G | -0.2518 | 0.047 | -0.1189 | 0.0278 |
| 3 | 23163749 | rs4634143 | T | C | 0.0743 | 0.0469 | 0.1024 | 0.0343 |
| 3 | 27537909 | rs13082711 | T | C | -0.2805 | 0.0546 | -0.1835 | 0.032 |
| 3 | 36964583 | rs4678915 | A | G | -0.0851 | 0.0427 | 0.03 | 0.0313 |
| 3 | 37539090 | rs267517 | A | G | -0.0898 | 0.042 | 0.0053 | 0.0308 |
| 3 | 38767315 | rs6801957 | T | C | 0.0706 | 0.0418 | -0.0941 | 0.0306 |
| 3 | 41107173 | rs6788984 | A | G | 0.168 | 0.061 | 0.0388 | 0.0447 |
| 3 | 41912651 | rs9815354 | A | G | -0.0628 | 0.0629 | 0.3264 | 0.0371 |
| 3 | 44858131 | rs141979279 | T | C | 0.2648 | 0.0963 | -0.0948 | 0.0702 |
| 3 | 46861939 | rs113134141 | A | G | -0.1352 | 0.0687 | -0.1451 | 0.0504 |
| 3 | 48197614 | rs6797587 | A | G | -0.28 | 0.0495 | -0.2209 | 0.0291 |
| 3 | 49913705 | rs36022378 | T | C | -0.1561 | 0.06 | -0.1144 | 0.0353 |
| 3 | 52558008 | rs13303 | T | C | -0.1095 | 0.0473 | -0.0153 | 0.0278 |
| 3 | 53635595 | rs9810888 | T | G | -0.1507 | 0.0464 | -0.128 | 0.0273 |
| 3 | 56726646 | rs9827472 | T | C | -0.1639 | 0.0485 | -0.1426 | 0.0285 |
| 3 | 57706503 | rs12486605 | T | C | -0.0278 | 0.0421 | -0.122 | 0.0308 |
| 3 | 63856870 | rs3774702 | A | G | 0.098 | 0.0615 | 0.1692 | 0.0361 |
| 3 | 64710253 | rs918466 | A | G | -0.1387 | 0.0477 | -0.1637 | 0.0281 |
| 3 | 66427029 | rs7630745 | T | C | 0.1319 | 0.0438 | 0.009 | 0.0321 |
| 3 | 73260545 | rs729448 | A | G | -0.0769 | 0.0465 | 0.0029 | 0.0274 |
| 3 | 74710462 | rs9857362 | A | C | 0.1104 | 0.0418 | 0.011 | 0.0306 |
| 3 | 85656311 | rs1375564 | T | C | 0.1884 | 0.0439 | 0.0441 | 0.0322 |
| 3 | 99839106 | rs9860290 | A | G | -0.1488 | 0.0529 | -0.0168 | 0.0388 |
| 3 | 111500002 | rs28675079 | A | G | -0.2597 | 0.0583 | -0.1657 | 0.0344 |
| 3 | 114461208 | rs1882289 | A | G | -0.2289 | 0.064 | -0.141 | 0.0469 |
| 3 | 123049938 | rs6806529 | A | C | 0.2028 | 0.0477 | 0.0673 | 0.028 |
| 3 | 124557643 | rs6438857 | T | C | 0.0977 | 0.0427 | 0.0089 | 0.0313 |
| 3 | 128201889 | rs62270945 | T | C | 0.1575 | 0.1581 | -0.2206 | 0.0911 |
| 3 | 132780356 | rs9875380 | T | C | -0.1456 | 0.0421 | -0.0663 | 0.0308 |
| 3 | 133959552 | rs6783086 | T | C | 0.257 | 0.0468 | 0.146 | 0.0275 |
| 3 | 138119952 | rs2306374 | T | C | -0.2268 | 0.0628 | -0.1699 | 0.037 |
| 3 | 141134818 | rs16851397 | A | G | -0.5907 | 0.1135 | -0.4 | 0.0664 |
| 3 | 142631909 | rs62278541 | A | G | 0.1037 | 0.0441 | -0.0265 | 0.0323 |
| 3 | 149524692 | rs6772704 | A | C | -0.1138 | 0.0501 | 0.0019 | 0.0366 |
| 3 | 154707967 | rs143112823 | A | G | -0.4019 | 0.0949 | -0.1784 | 0.0552 |
| 3 | 158316726 | rs78151625 | T | C | -0.1451 | 0.0562 | -0.0583 | 0.0411 |
| 3 | 169100886 | rs419076 | T | C | 0.3947 | 0.0457 | 0.25 | 0.0269 |
| 3 | 171995605 | rs4894535 | T | C | 0.1221 | 0.0631 | -0.0105 | 0.0371 |
| 3 | 176927949 | rs73171158 | T | C | -0.2109 | 0.0499 | -0.0839 | 0.0293 |
| 3 | 179169230 | rs7611674 | T | G | 0.1299 | 0.0599 | 0.1549 | 0.0353 |
| 3 | 183435713 | rs262986 | A | G | -0.2288 | 0.0468 | -0.0673 | 0.0275 |
| 3 | 194299967 | rs1706003 | T | G | 0.096 | 0.0429 | 0.1576 | 0.0314 |
| 3 | 197070959 | rs6777317 | A | G | 0.1642 | 0.045 | 0.0855 | 0.033 |
| 4 | 1254930 | rs1250129 | A | G | -0.1673 | 0.0733 | 0.0096 | 0.043 |
| 4 | 2165493 | rs55829085 | A | C | -0.226 | 0.1039 | -0.219 | 0.0758 |
| 4 | 3451109 | rs2498323 | A | G | 0.1963 | 0.0703 | -0.1273 | 0.0518 |
| 4 | 15356795 | rs13122790 | A | G | 0.0677 | 0.0486 | -0.0515 | 0.0355 |
| 4 | 16032948 | rs11730129 | T | C | -0.2053 | 0.0561 | 0.0102 | 0.033 |
| 4 | 18008232 | rs2610990 | A | G | -0.1915 | 0.0484 | -0.0165 | 0.0355 |
| 4 | 38387395 | rs2291435 | T | C | -0.2419 | 0.0463 | -0.1349 | 0.0272 |
| 4 | 46595623 | rs12511987 | T | G | -0.1817 | 0.0551 | -0.0876 | 0.0402 |

| | | | | | | | | |
|---|---|---|---|---|---|---|---|---|
| 4 | 54799245 | rs871606 | T | C | 0.3598 | 0.076 | -0.0896 | 0.0448 |
| 4 | 57943153 | rs1718845 | A | G | -0.0131 | 0.0453 | -0.0907 | 0.0332 |
| 4 | 77414144 | rs10008637 | T | C | 0.1355 | 0.0415 | 0.0972 | 0.0304 |
| 4 | 81164723 | rs1458038 | T | C | 0.6612 | 0.0507 | 0.4107 | 0.0298 |
| 4 | 83925895 | rs6823199 | T | C | 0.2267 | 0.0503 | 0.0734 | 0.0369 |
| 4 | 86715670 | rs2014912 | T | C | 0.5122 | 0.0644 | 0.1492 | 0.0379 |
| 4 | 89750668 | rs13149209 | T | C | 0.196 | 0.0496 | 0.054 | 0.0363 |
| 4 | 95938386 | rs1347345 | A | G | -0.1765 | 0.0433 | -0.1153 | 0.0317 |
| 4 | 102435265 | rs17248480 | A | G | -0.5467 | 0.1312 | -0.4718 | 0.0964 |
| 4 | 103188709 | rs13107325 | T | C | -0.8783 | 0.0947 | -0.6042 | 0.0556 |
| 4 | 103769304 | rs223361 | T | C | -0.043 | 0.0456 | 0.0942 | 0.0334 |
| 4 | 106109381 | rs4699165 | A | G | 0.083 | 0.0483 | -0.0456 | 0.0284 |
| 4 | 109017528 | rs7694643 | A | G | -0.183 | 0.0432 | -0.0874 | 0.0317 |
| 4 | 111381638 | rs6825911 | T | C | -0.2376 | 0.0576 | -0.1712 | 0.034 |
| 4 | 119958809 | rs4834735 | T | C | 0.1927 | 0.0676 | 0.104 | 0.0397 |
| 4 | 120509279 | rs66887589 | T | C | -0.1812 | 0.0463 | -0.1251 | 0.0272 |
| 4 | 138464842 | rs7439567 | T | C | 0.1998 | 0.0432 | 0.1216 | 0.0316 |
| 4 | 146821725 | rs4835266 | T | C | 0.0766 | 0.0445 | -0.0409 | 0.0326 |
| 4 | 148400256 | rs10305838 | T | C | -0.117 | 0.0607 | 0.0207 | 0.0445 |
| 4 | 151295085 | rs6823767 | T | C | -0.1566 | 0.0528 | -0.0839 | 0.031 |
| 4 | 156645513 | rs13139571 | A | C | -0.2959 | 0.0538 | -0.2307 | 0.0316 |
| 4 | 157678511 | rs17035181 | T | G | 0.271 | 0.0593 | 0.1245 | 0.0434 |
| 4 | 169688000 | rs869396 | A | C | -0.1803 | 0.0466 | -0.0079 | 0.0274 |
| 5 | 361148 | rs4957026 | A | G | 0.2214 | 0.0497 | 0.1048 | 0.0292 |
| 5 | 1279790 | rs10069690 | T | C | 0.2151 | 0.0468 | 0.118 | 0.0343 |
| 5 | 3706050 | rs954767 | A | C | -0.1658 | 0.0465 | -0.1074 | 0.034 |
| 5 | 32815028 | rs1173771 | A | G | -0.5227 | 0.0468 | -0.2582 | 0.0276 |
| 5 | 43824677 | rs7710854 | A | G | 0.254 | 0.0663 | 0.0439 | 0.0485 |
| 5 | 52135543 | rs6867399 | A | C | 0.0834 | 0.0542 | -0.0585 | 0.0396 |
| 5 | 55868097 | rs13179413 | T | C | 0.1656 | 0.0492 | 0.1214 | 0.0361 |
| 5 | 57095011 | rs12515541 | T | G | 0.2175 | 0.0431 | 0.0939 | 0.0316 |
| 5 | 57754005 | rs1848510 | A | G | 0.1129 | 0.0486 | 0.0736 | 0.0286 |
| 5 | 61553881 | rs10062049 | T | C | 0.1849 | 0.0611 | 0.1821 | 0.0447 |
| 5 | 65662133 | rs3121685 | T | C | -0.1302 | 0.0422 | -0.0592 | 0.0309 |
| 5 | 66291370 | rs4286632 | A | G | 0.148 | 0.0471 | 0.1548 | 0.0345 |
| 5 | 68007803 | rs246973 | T | C | 0.1721 | 0.0499 | 0.0796 | 0.0365 |
| 5 | 71506529 | rs72761109 | T | C | 0.0874 | 0.0474 | -0.0216 | 0.0347 |
| 5 | 72654304 | rs4443403 | T | C | -0.1725 | 0.0687 | -0.0286 | 0.0502 |
| 5 | 75038431 | rs10078021 | T | G | -0.0619 | 0.0489 | -0.1218 | 0.0288 |
| 5 | 77837789 | rs10057188 | A | G | -0.1942 | 0.0475 | -0.0671 | 0.028 |
| 5 | 87514515 | rs10059921 | T | G | -0.3732 | 0.0919 | -0.219 | 0.0542 |
| 5 | 89484911 | rs62380354 | A | C | 0.2183 | 0.0833 | 0.1982 | 0.0484 |
| 5 | 96174186 | rs709668 | A | G | -0.1743 | 0.0524 | -0.131 | 0.0384 |
| 5 | 97953719 | rs1871190 | T | G | 0.1658 | 0.0495 | 0.1062 | 0.0291 |
| 5 | 107458637 | rs286809 | A | G | -0.181 | 0.0563 | -0.0172 | 0.0412 |
| 5 | 108113740 | rs79409628 | T | G | -0.265 | 0.0744 | 0.0957 | 0.0544 |
| 5 | 114390121 | rs10077885 | A | C | -0.2465 | 0.0484 | -0.1321 | 0.0285 |
| 5 | 119781569 | rs1432457 | A | G | -0.1559 | 0.0461 | -0.0368 | 0.0338 |
| 5 | 121194226 | rs9885577 | T | C | 0.1325 | 0.0446 | 0.0433 | 0.0327 |
| 5 | 122476457 | rs13359291 | A | G | 0.4005 | 0.062 | 0.1614 | 0.0366 |
| 5 | 123136656 | rs6891344 | A | G | 0.284 | 0.0599 | 0.2182 | 0.0354 |
| 5 | 127868199 | rs6595838 | A | G | 0.2361 | 0.0507 | 0.1216 | 0.0297 |
| 5 | 131784393 | rs12521868 | T | G | -0.123 | 0.0476 | -0.1463 | 0.028 |
| 5 | 132397351 | rs55747751 | A | G | -0.1589 | 0.0792 | -0.1912 | 0.0581 |
| 5 | 140086677 | rs702395 | T | C | 0.115 | 0.0416 | 0.0223 | 0.0305 |
| 5 | 147696018 | rs2400509 | A | G | -0.1076 | 0.0524 | 1.00E-04 | 0.0308 |
| 5 | 148391140 | rs9687065 | A | G | 0.2501 | 0.059 | 0.1789 | 0.0348 |
| 5 | 157845402 | rs11953630 | T | C | -0.4463 | 0.0501 | -0.2185 | 0.0295 |
| 5 | 172192350 | rs114503346 | T | C | -0.1495 | 0.1198 | -0.276 | 0.0698 |
| 5 | 176731452 | rs28362590 | T | G | 0.1061 | 0.0548 | 0.1196 | 0.0322 |
| 5 | 179411477 | rs12153395 | A | G | -0.2602 | 0.0764 | -0.0841 | 0.0449 |
| 6 | 1613686 | rs2745599 | A | G | 0.1464 | 0.0418 | 0.055 | 0.0306 |
| 6 | 7211818 | rs1334576 | A | G | -0.0736 | 0.0416 | 0.0436 | 0.0305 |
| 6 | 12295987 | rs1630736 | T | C | -0.1846 | 0.042 | -0.0401 | 0.0308 |
| 6 | 12903957 | rs9349379 | A | G | 0.2624 | 0.0485 | 0.0727 | 0.0285 |
| 6 | 19028623 | rs12216497 | T | C | 0.1211 | 0.0461 | 0.003 | 0.0272 |
| 6 | 20686996 | rs9368222 | A | C | 0.2337 | 0.0462 | 0.0448 | 0.0338 |

| | | | | | | | | |
|---|---|---|---|---|---|---|---|---|
| 6 | 22130601 | rs6911827 | T | C | 0.152 | 0.0473 | 0.12 | 0.0279 |
| 6 | 31708147 | rs409558 | T | C | 0.4456 | 0.0637 | 0.1347 | 0.0377 |
| 6 | 34244132 | rs115245297 | T | C | -0.5045 | 0.1111 | -0.3259 | 0.0816 |
| 6 | 43308363 | rs1563788 | T | C | 0.3062 | 0.0501 | 0.1301 | 0.0295 |
| 6 | 43809802 | rs9472135 | T | C | 0.0094 | 0.0455 | 0.1126 | 0.0333 |
| 6 | 50683009 | rs78648104 | T | C | -0.3571 | 0.083 | -0.1824 | 0.049 |
| 6 | 51832494 | rs13205180 | T | C | 0.041 | 0.0464 | 0.1195 | 0.0273 |
| 6 | 53994626 | rs631441 | T | G | -0.0462 | 0.0449 | 0.0537 | 0.0329 |
| 6 | 56102780 | rs1925153 | T | C | -0.0511 | 0.0492 | 0.0724 | 0.0288 |
| 6 | 72206620 | rs504691 | A | C | -0.0098 | 0.047 | -0.0817 | 0.0277 |
| 6 | 73657714 | rs12195276 | T | C | -0.1819 | 0.0471 | 0.009 | 0.0345 |
| 6 | 79655477 | rs10943605 | A | G | 0.1747 | 0.0462 | 0.1552 | 0.0272 |
| 6 | 80818531 | rs7753695 | T | C | 0.1176 | 0.0476 | 0.098 | 0.028 |
| 6 | 82281417 | rs9449350 | T | C | -0.2333 | 0.0488 | -0.1346 | 0.0287 |
| 6 | 85283253 | rs60255247 | A | C | 0.0788 | 0.0757 | -0.0474 | 0.0445 |
| 6 | 96885405 | rs35410524 | T | C | 0.2999 | 0.0588 | 0.1412 | 0.0347 |
| 6 | 109013930 | rs9486916 | T | C | 0.2572 | 0.058 | 0.066 | 0.0341 |
| 6 | 117523671 | rs2693560 | A | G | -0.1978 | 0.0448 | -0.1391 | 0.0329 |
| 6 | 118026126 | rs2498586 | T | C | -0.1617 | 0.0568 | -0.0919 | 0.0416 |
| 6 | 119113317 | rs9401090 | T | C | 0.1762 | 0.0494 | 0.0842 | 0.0362 |
| 6 | 121781390 | rs11154027 | T | C | -0.009 | 0.0475 | -0.0655 | 0.0279 |
| 6 | 122287990 | rs6925750 | T | C | 0.0726 | 0.0637 | -0.0916 | 0.0466 |
| 6 | 126228512 | rs10782230 | A | G | 0.1776 | 0.0425 | 0.0548 | 0.0311 |
| 6 | 127115454 | rs13209747 | T | C | 0.4117 | 0.0466 | 0.2542 | 0.0274 |
| 6 | 131311909 | rs9885632 | T | C | 0.1582 | 0.0486 | 0.0261 | 0.0357 |
| 6 | 135119089 | rs4896104 | T | C | -0.0999 | 0.0433 | 0.0606 | 0.0317 |
| 6 | 139835689 | rs668459 | T | C | -0.1357 | 0.0419 | -0.1487 | 0.0307 |
| 6 | 140383733 | rs7763294 | T | G | -0.1418 | 0.045 | -0.046 | 0.0329 |
| 6 | 147713764 | rs7765526 | A | G | 0.2317 | 0.047 | 0.1251 | 0.0277 |
| 6 | 152398505 | rs9479200 | A | G | -0.0985 | 0.0709 | 0.1494 | 0.0418 |
| 6 | 153427265 | rs9479509 | A | G | -0.1242 | 0.0497 | -0.0953 | 0.0293 |
| 6 | 154418759 | rs598682 | A | G | -0.155 | 0.0476 | -0.1398 | 0.0349 |
| 6 | 160769423 | rs555754 | A | G | -0.1895 | 0.0459 | -0.0505 | 0.027 |
| 6 | 161712235 | rs9456648 | T | C | -0.2089 | 0.0492 | -0.1459 | 0.029 |
| 6 | 169587103 | rs1322639 | A | G | 0.0888 | 0.0557 | -0.1312 | 0.0328 |
| 7 | 1195692 | rs73033340 | A | G | 0.3412 | 0.1287 | 0.2476 | 0.092 |
| 7 | 1966831 | rs6959688 | A | G | -0.1323 | 0.0437 | -0.0421 | 0.032 |
| 7 | 2512545 | rs2969070 | A | G | -0.2752 | 0.0478 | -0.198 | 0.0281 |
| 7 | 4669949 | rs73049928 | A | G | -0.1323 | 0.0541 | -0.1084 | 0.0396 |
| 7 | 7290732 | rs1468520 | A | G | -0.1466 | 0.0615 | -0.1269 | 0.0362 |
| 7 | 14375977 | rs13240040 | A | G | 0.1457 | 0.0518 | 0.1161 | 0.0304 |
| 7 | 19049388 | rs2107595 | A | G | 0.3022 | 0.0623 | -0.0518 | 0.0367 |
| 7 | 25871109 | rs1055144 | T | C | 0.108 | 0.0582 | 2.00E-04 | 0.0342 |
| 7 | 28142088 | rs10274928 | A | G | 0.1505 | 0.0411 | 0.0515 | 0.0302 |
| 7 | 28658522 | rs917275 | A | G | -0.1133 | 0.0478 | 0.0351 | 0.0282 |
| 7 | 35467896 | rs342989 | A | G | 0.0588 | 0.0557 | 0.1161 | 0.0328 |
| 7 | 36225818 | rs2052263 | A | G | 0.2349 | 0.0787 | 0.0171 | 0.0458 |
| 7 | 40447971 | rs76206723 | A | G | -0.3356 | 0.0744 | -0.0328 | 0.0438 |
| 7 | 44240407 | rs1004558 | T | C | 0.1803 | 0.0552 | -0.0237 | 0.0406 |
| 7 | 45036785 | rs73105827 | T | G | -0.1782 | 0.0864 | -0.1437 | 0.0508 |
| 7 | 46008110 | rs11977526 | A | G | -0.2535 | 0.0484 | 0.0511 | 0.0285 |
| 7 | 46554358 | rs71543920 | T | G | -0.2962 | 0.0927 | 0.0359 | 0.068 |
| 7 | 47548893 | rs12668436 | T | C | -0.1554 | 0.0534 | -0.0026 | 0.0314 |
| 7 | 50915776 | rs17454517 | A | G | 0.0718 | 0.0466 | 0.1276 | 0.0274 |
| 7 | 69769369 | rs2222544 | T | C | -0.0333 | 0.0482 | -0.094 | 0.0353 |
| 7 | 73491212 | rs1091811 | A | G | -0.0775 | 0.0625 | 0.0394 | 0.0367 |
| 7 | 74107374 | rs34324971 | A | G | 0.0828 | 0.0532 | 0.1341 | 0.039 |
| 7 | 75097488 | rs6963105 | A | G | -0.1548 | 0.046 | -0.0939 | 0.0336 |
| 7 | 77572461 | rs848445 | T | C | -0.196 | 0.049 | -0.0322 | 0.0359 |
| 7 | 89805241 | rs11770630 | T | C | 0.0556 | 0.0413 | -0.1108 | 0.0303 |
| 7 | 90449362 | rs10245696 | A | C | 0.105 | 0.0468 | -0.0135 | 0.0275 |
| 7 | 92264410 | rs2282978 | T | C | 0.0704 | 0.0487 | -0.0518 | 0.0288 |
| 7 | 96461649 | rs1947228 | T | C | -0.0432 | 0.0414 | -0.146 | 0.0304 |
| 7 | 100467700 | rs12705090 | T | C | -0.077 | 0.0525 | 0.1183 | 0.0384 |
| 7 | 106411858 | rs17477177 | T | C | -0.5642 | 0.0564 | -0.0153 | 0.0332 |
| 7 | 107019947 | rs115172170 | T | C | -0.3593 | 0.0887 | -0.0856 | 0.0648 |
| 7 | 116198621 | rs1997571 | A | G | -0.0912 | 0.042 | 0.0414 | 0.0308 |

| | | | | | | | | |
|---|---|---|---|---|---|---|---|---|
| 7 | 128573967 | rs4728142 | A | G | -0.2155 | 0.0467 | -0.0752 | 0.0275 |
| 7 | 129663496 | rs11556924 | T | C | -0.2487 | 0.0498 | -0.142 | 0.0293 |
| 7 | 130432469 | rs34072724 | A | G | -0.1735 | 0.0412 | -0.0784 | 0.0302 |
| 7 | 131059056 | rs13238550 | A | G | 0.1695 | 0.0472 | 0.0892 | 0.0277 |
| 7 | 140238048 | rs12703989 | A | G | 0.1501 | 0.0436 | 0.0621 | 0.0319 |
| 7 | 149474622 | rs73727605 | A | G | 0.3034 | 0.0994 | 0.0601 | 0.0585 |
| 7 | 150050111 | rs11771693 | A | G | 0.1368 | 0.0455 | 0.0328 | 0.0333 |
| 7 | 150690176 | rs3918226 | T | C | 0.5473 | 0.091 | 0.5454 | 0.0534 |
| 7 | 150704843 | rs891511 | A | G | -0.2529 | 0.0521 | -0.2109 | 0.0306 |
| 7 | 151415041 | rs10224002 | A | G | -0.2375 | 0.0525 | -0.1368 | 0.031 |
| 7 | 156311745 | rs9638084 | A | G | 0.1369 | 0.0429 | 0.0959 | 0.0314 |
| 8 | 1721090 | rs4875958 | A | G | 0.2209 | 0.0515 | 0.0898 | 0.0304 |
| 8 | 8503700 | rs61040371 | T | C | 0.191 | 0.0475 | 0.0801 | 0.0279 |
| 8 | 9730663 | rs62491354 | A | G | 0.1582 | 0.0608 | 0.103 | 0.0445 |
| 8 | 10268736 | rs1986971 | A | G | 0.1991 | 0.0478 | 0.0754 | 0.035 |
| 8 | 11433909 | rs2898290 | T | C | 0.3419 | 0.0466 | 0.1324 | 0.0274 |
| 8 | 17427186 | rs75902664 | A | G | -0.1995 | 0.171 | -0.3143 | 0.0989 |
| 8 | 22428708 | rs1047030 | A | G | 0.1866 | 0.0591 | 0.1383 | 0.0431 |
| 8 | 23400615 | rs62503324 | T | C | 0.1569 | 0.0474 | 0.1635 | 0.0347 |
| 8 | 25900675 | rs6557876 | T | C | -0.3667 | 0.0533 | -0.1966 | 0.0314 |
| 8 | 26445194 | rs17321041 | T | C | 0.3664 | 0.0971 | 0.2722 | 0.0569 |
| 8 | 30288272 | rs2979470 | T | C | 0.2114 | 0.046 | 0.1003 | 0.027 |
| 8 | 33309993 | rs7845722 | A | G | -0.1209 | 0.0467 | -0.0068 | 0.0275 |
| 8 | 38130025 | rs1906672 | A | G | 0.2227 | 0.0499 | 0.0752 | 0.0366 |
| 8 | 42324765 | rs2978456 | T | C | -0.1069 | 0.0498 | -0.0121 | 0.0291 |
| 8 | 51947549 | rs4873492 | T | C | 0.2131 | 0.0584 | 0.1059 | 0.0427 |
| 8 | 60535824 | rs6996733 | T | C | 0.1484 | 0.0568 | 0.0656 | 0.0416 |
| 8 | 64501744 | rs2354862 | A | C | 0.2139 | 0.0485 | 0.0939 | 0.0285 |
| 8 | 68920135 | rs13253358 | T | C | 0.1945 | 0.0504 | 0.123 | 0.0296 |
| 8 | 76054904 | rs1350100 | A | G | 0.0382 | 0.043 | -0.0591 | 0.0315 |
| 8 | 76591880 | rs1449544 | A | C | 0.2215 | 0.0459 | 0.0368 | 0.027 |
| 8 | 81393697 | rs72688070 | T | C | -0.187 | 0.0549 | -0.1213 | 0.0403 |
| 8 | 82814156 | rs56345595 | A | G | 0.1538 | 0.043 | 0.0614 | 0.0315 |
| 8 | 92149429 | rs7009170 | T | C | -0.0775 | 0.0451 | 0.0529 | 0.0331 |
| 8 | 92769569 | rs62526122 | A | G | 0.1739 | 0.0557 | 0.1116 | 0.0326 |
| 8 | 95969257 | rs4582532 | A | G | -0.091 | 0.0411 | 0.0126 | 0.0301 |
| 8 | 101676675 | rs2978098 | A | C | 0.1483 | 0.0474 | 0.1223 | 0.0279 |
| 8 | 102750597 | rs142449193 | T | C | -0.4354 | 0.112 | -0.2426 | 0.0663 |
| 8 | 103883630 | rs2513877 | A | G | -0.1413 | 0.0576 | -0.1107 | 0.034 |
| 8 | 105966258 | rs35783704 | A | G | -0.5219 | 0.0773 | -0.241 | 0.0454 |
| 8 | 110107161 | rs28499085 | A | G | 0.1426 | 0.0486 | 0.0319 | 0.0355 |
| 8 | 116959837 | rs2205260 | A | C | 0.0555 | 0.0559 | -0.085 | 0.0409 |
| 8 | 120435812 | rs2071518 | T | C | 0.2181 | 0.0524 | -0.1409 | 0.0308 |
| 8 | 126520544 | rs62523863 | A | G | 0.1643 | 0.0502 | 0.0686 | 0.0368 |
| 8 | 129483956 | rs4598218 | T | C | 0.1578 | 0.0435 | 0.0308 | 0.0318 |
| 8 | 135612745 | rs894344 | A | G | -0.1455 | 0.0465 | -0.0915 | 0.0274 |
| 8 | 141060027 | rs4454254 | A | G | -0.2213 | 0.0475 | -0.0186 | 0.028 |
| 8 | 141858620 | rs10087782 | T | C | 0.14 | 0.0418 | 0.0872 | 0.0306 |
| 8 | 142367087 | rs34591516 | T | C | 0.5607 | 0.1067 | 0.2869 | 0.0632 |
| 8 | 143312933 | rs4129585 | A | C | 0.1541 | 0.0414 | 0.0872 | 0.0303 |
| 8 | 144060955 | rs62524579 | A | G | -0.1811 | 0.0533 | -0.1504 | 0.0309 |
| 8 | 144981488 | rs56233017 | A | G | -0.1275 | 0.1115 | -0.2413 | 0.0815 |
| 9 | 753648 | rs60191654 | A | G | -0.2311 | 0.0584 | -0.1328 | 0.0348 |
| 9 | 2493751 | rs12216886 | T | G | 0.1226 | 0.057 | 0.0949 | 0.0341 |
| 9 | 9350706 | rs1332813 | T | C | 0.1453 | 0.046 | 0.132 | 0.0337 |
| 9 | 10594635 | rs35287509 | T | C | -0.2083 | 0.0489 | -0.1218 | 0.0292 |
| 9 | 16872323 | rs11789875 | A | G | 0.1265 | 0.0596 | -0.0413 | 0.0436 |
| 9 | 19057551 | rs4977492 | T | C | -0.0939 | 0.0444 | 0.0177 | 0.0326 |
| 9 | 21801530 | rs4364717 | A | G | -0.1065 | 0.0455 | -0.0855 | 0.0272 |
| 9 | 22942770 | rs9886665 | T | C | 0.1887 | 0.0519 | 0.1087 | 0.031 |
| 9 | 34223553 | rs4553000 | T | C | -0.1627 | 0.0452 | -0.0405 | 0.027 |
| 9 | 35906471 | rs76452347 | T | C | -0.172 | 0.0624 | -0.1691 | 0.0372 |
| 9 | 85128518 | rs11139596 | T | G | -0.1876 | 0.0574 | -0.0031 | 0.0342 |
| 9 | 89888472 | rs11141731 | T | C | -0.1955 | 0.0549 | -0.1315 | 0.0328 |
| 9 | 101739709 | rs10988442 | A | G | 0.1174 | 0.0428 | -0.056 | 0.0313 |
| 9 | 112358150 | rs7043304 | T | C | 0.0488 | 0.0657 | 0.1218 | 0.0392 |
| 9 | 113169775 | rs111245230 | T | C | -0.6917 | 0.1299 | -0.4 | 0.0777 |

| | | | | | | | | |
|---|---|---|---|---|---|---|---|---|
| 9 | 116696625 | rs13290326 | T | C | -0.162 | 0.0428 | -0.0461 | 0.0313 |
| 9 | 118534500 | rs10982910 | T | G | -0.2591 | 0.0725 | -0.0164 | 0.0532 |
| 9 | 119312256 | rs1861881 | T | G | 0.0717 | 0.0486 | 0.0862 | 0.029 |
| 9 | 123640500 | rs1953126 | T | C | 0.1838 | 0.0478 | 0.0141 | 0.0286 |
| 9 | 125755571 | rs10818775 | T | C | -0.2731 | 0.0696 | -0.0256 | 0.0416 |
| 9 | 127900996 | rs72765298 | T | C | -0.3477 | 0.0726 | -0.0413 | 0.0432 |
| 9 | 128498594 | rs7023828 | T | C | -0.2125 | 0.0427 | -0.0369 | 0.0312 |
| 9 | 130309028 | rs1891730 | T | C | -0.1733 | 0.0441 | -0.1081 | 0.0323 |
| 9 | 131210410 | rs7869756 | A | G | -0.0479 | 0.0552 | 0.0769 | 0.0404 |
| 9 | 131940019 | rs184457 | A | G | -0.1527 | 0.0494 | -0.0698 | 0.0361 |
| 9 | 136137065 | rs687621 | A | G | 0.0979 | 0.0484 | 0.1327 | 0.029 |
| 9 | 136522274 | rs6271 | T | C | -0.4339 | 0.102 | -0.3647 | 0.0613 |
| 9 | 139520789 | rs11145807 | A | G | 0.1532 | 0.0522 | 0.1429 | 0.0306 |
| 10 | 5804865 | rs56352451 | T | C | 0.1563 | 0.0603 | 0.0143 | 0.0442 |
| 10 | 10876943 | rs36006409 | T | G | -0.2507 | 0.0553 | -0.0313 | 0.0405 |
| 10 | 13523937 | rs10906391 | T | C | 0.1366 | 0.0452 | 0.0976 | 0.0331 |
| 10 | 18707448 | rs1813353 | T | C | 0.4324 | 0.0482 | 0.2772 | 0.0284 |
| 10 | 20531420 | rs72795925 | T | C | 0.0842 | 0.0498 | -0.0388 | 0.0365 |
| 10 | 21037294 | rs10732433 | T | C | 0.1051 | 0.0423 | -0.0558 | 0.031 |
| 10 | 28924901 | rs1265842 | T | C | 0.1634 | 0.0464 | 0.1444 | 0.0273 |
| 10 | 30317073 | rs9337951 | A | G | 0.0928 | 0.0528 | -0.1128 | 0.031 |
| 10 | 32082658 | rs10826995 | T | C | -0.0896 | 0.0514 | 0.0207 | 0.0302 |
| 10 | 32590362 | rs76164690 | T | G | -0.1393 | 0.0682 | -0.1702 | 0.0398 |
| 10 | 45273079 | rs2246438 | A | G | -0.0217 | 0.0512 | -0.0714 | 0.0301 |
| 10 | 48411796 | rs34130368 | T | G | -0.2205 | 0.0647 | -0.1209 | 0.0476 |
| 10 | 62390726 | rs10761530 | T | C | 0.2533 | 0.0455 | 0.1421 | 0.0268 |
| 10 | 63524591 | rs1530440 | T | C | -0.4983 | 0.0587 | -0.3586 | 0.0345 |
| 10 | 65335315 | rs7090758 | T | C | -0.1248 | 0.0426 | -0.0755 | 0.0312 |
| 10 | 69350563 | rs7914287 | T | C | -0.1487 | 0.0559 | -0.0017 | 0.0329 |
| 10 | 69855363 | rs10823136 | T | C | -0.3274 | 0.0857 | -0.1161 | 0.0508 |
| 10 | 70404159 | rs10998362 | T | C | 0.0609 | 0.0531 | -0.0543 | 0.0311 |
| 10 | 74751579 | rs12572586 | T | C | -0.3177 | 0.0874 | -0.2126 | 0.0639 |
| 10 | 82215288 | rs10887914 | T | C | 0.1523 | 0.0412 | 0.0465 | 0.0302 |
| 10 | 89681688 | rs77413490 | T | G | 0.522 | 0.1091 | 0.1808 | 0.0795 |
| 10 | 94468685 | rs11187142 | T | C | 0.298 | 0.0763 | 0.0721 | 0.0447 |
| 10 | 95895940 | rs932764 | A | G | -0.3654 | 0.0467 | -0.148 | 0.0275 |
| 10 | 96563757 | rs4494250 | A | G | 0.2561 | 0.0486 | 0.1767 | 0.0286 |
| 10 | 102075479 | rs603424 | A | G | 0.1454 | 0.0574 | 0.1518 | 0.0421 |
| 10 | 102604514 | rs112184198 | A | G | -0.5331 | 0.0761 | -0.3003 | 0.0448 |
| 10 | 103115345 | rs72847884 | A | G | 0.2555 | 0.1182 | 0.3526 | 0.0691 |
| 10 | 103702763 | rs11191156 | A | G | -0.0829 | 0.0484 | 0.0237 | 0.0285 |
| 10 | 104846178 | rs11191548 | T | C | 1.0233 | 0.0818 | 0.4546 | 0.0482 |
| 10 | 105677897 | rs4387287 | A | C | 0.1953 | 0.0624 | 0.1671 | 0.0368 |
| 10 | 106894942 | rs191784289 | T | C | 1.1113 | 0.2324 | 0.802 | 0.135 |
| 10 | 111965826 | rs111777102 | T | C | 0.1692 | 0.0933 | 0.1923 | 0.0552 |
| 10 | 114754071 | rs34872471 | T | C | -0.1796 | 0.0509 | -0.0066 | 0.03 |
| 10 | 115781527 | rs2782980 | T | C | -0.3388 | 0.0521 | -0.2424 | 0.0307 |
| 10 | 118523933 | rs11197813 | A | G | -0.1765 | 0.046 | -0.0517 | 0.0338 |
| 10 | 121433675 | rs72842207 | T | C | -0.0249 | 0.0509 | -0.211 | 0.0373 |
| 10 | 122968964 | rs11592107 | A | G | 0.2721 | 0.0495 | 0.112 | 0.0292 |
| 10 | 124235226 | rs72834453 | T | G | -0.2378 | 0.0712 | -0.0932 | 0.0421 |
| 10 | 126712781 | rs4411245 | A | G | 0.1204 | 0.0457 | 0.1317 | 0.0335 |
| 10 | 134459388 | rs1133400 | A | G | -0.193 | 0.051 | -0.051 | 0.0374 |
| 11 | 828916 | rs7126805 | A | G | 0.1195 | 0.0555 | -0.0099 | 0.0326 |
| 11 | 1905292 | rs661348 | T | C | -0.3417 | 0.0502 | -0.1612 | 0.0293 |
| 11 | 4673788 | rs17224476 | A | G | 0.1437 | 0.0668 | 0.1389 | 0.0492 |
| 11 | 6289118 | rs2929184 | A | G | 0.1035 | 0.0528 | 0.1397 | 0.0386 |
| 11 | 8252853 | rs110419 | A | G | 0.0808 | 0.047 | 0.1167 | 0.0276 |
| 11 | 8774923 | rs10743086 | A | G | -0.14 | 0.0522 | -0.0665 | 0.0382 |
| 11 | 9762274 | rs360153 | T | C | -0.186 | 0.0423 | -0.1036 | 0.031 |
| 11 | 10350538 | rs7129220 | A | G | 0.3919 | 0.0724 | 0.1886 | 0.0426 |
| 11 | 13293905 | rs900145 | T | C | 0.1135 | 0.0499 | 0.1191 | 0.0294 |
| 11 | 16302939 | rs4757391 | T | C | -0.4921 | 0.0573 | -0.2997 | 0.0337 |
| 11 | 16902268 | rs381815 | T | C | 0.3644 | 0.0515 | 0.2043 | 0.0303 |
| 11 | 17409572 | rs5219 | T | C | 0.32 | 0.0471 | 0.1578 | 0.0278 |
| 11 | 22515533 | rs11026586 | A | G | 0.1523 | 0.0845 | 0.1672 | 0.0613 |
| 11 | 27728102 | rs11030119 | A | G | -0.1699 | 0.0509 | -0.1332 | 0.03 |

| | | | | | | | | |
|---|---|---|---|---|---|---|---|---|
| 11 | 28512458 | rs871004 | A | G | 0.1329 | 0.0447 | 0.0638 | 0.0327 |
| 11 | 30355707 | rs11031051 | A | C | -0.127 | 0.0445 | -0.0903 | 0.0326 |
| 11 | 31111810 | rs919045 | T | C | 0.0636 | 0.0473 | 0.0801 | 0.0279 |
| 11 | 32374199 | rs4922591 | T | C | -0.2058 | 0.0478 | -0.056 | 0.0281 |
| 11 | 34068037 | rs190194639 | T | C | 0.2211 | 0.0802 | 0.199 | 0.0584 |
| 11 | 45207851 | rs7480089 | A | G | -0.2334 | 0.0716 | -0.0146 | 0.0422 |
| 11 | 47461783 | rs7103648 | A | G | -0.3102 | 0.0474 | -0.1879 | 0.0279 |
| 11 | 47587452 | rs11537751 | T | C | 0.3936 | 0.1076 | 0.2114 | 0.0628 |
| 11 | 55113534 | rs75905900 | A | C | 0.328 | 0.0647 | 0.1726 | 0.0474 |
| 11 | 57496820 | rs11607056 | T | C | -0.1143 | 0.0437 | 0.0248 | 0.032 |
| 11 | 58207203 | rs11229457 | T | C | -0.2886 | 0.0563 | -0.1588 | 0.0332 |
| 11 | 61278246 | rs751984 | T | C | 0.4344 | 0.0738 | 0.3632 | 0.0434 |
| 11 | 63744609 | rs4980515 | T | C | 0.1722 | 0.0447 | 0.0748 | 0.0327 |
| 11 | 65408937 | rs3741378 | T | C | -0.4169 | 0.0696 | -0.1823 | 0.0409 |
| 11 | 69079707 | rs67330701 | T | C | -0.2769 | 0.0913 | -0.231 | 0.0534 |
| 11 | 70005641 | rs875106 | A | G | 0.0636 | 0.0422 | -0.0858 | 0.0309 |
| 11 | 72006086 | rs504217 | T | C | 0.462 | 0.0887 | 0.2777 | 0.0523 |
| 11 | 73068571 | rs2298807 | T | C | 0.1428 | 0.0556 | 0.1071 | 0.0326 |
| 11 | 74374950 | rs4420291 | A | G | 0.0716 | 0.0419 | 0.0832 | 0.0307 |
| 11 | 76125330 | rs7927515 | A | C | 0.1705 | 0.0488 | 0.0909 | 0.0288 |
| 11 | 77940075 | rs2450128 | A | G | -0.1589 | 0.0634 | -0.1535 | 0.0372 |
| 11 | 89224453 | rs2289125 | A | C | -0.1944 | 0.0582 | 0.079 | 0.0343 |
| 11 | 101100768 | rs61892344 | T | C | -0.1007 | 0.0554 | -0.1129 | 0.0405 |
| 11 | 102077200 | rs12807220 | A | G | -0.1028 | 0.0442 | 0.0699 | 0.0324 |
| 11 | 107096777 | rs4754196 | A | G | -0.1756 | 0.0415 | -0.1026 | 0.0304 |
| 11 | 116772441 | rs1076485 | T | C | 0.2665 | 0.0606 | 0.0826 | 0.0442 |
| 11 | 117283676 | rs8258 | T | C | 0.0559 | 0.0478 | -0.0868 | 0.0281 |
| 11 | 122521123 | rs12574332 | T | C | 0.2026 | 0.0703 | 0.1812 | 0.0413 |
| 12 | 2521579 | rs55935819 | A | G | 0.2044 | 0.0475 | 0.1222 | 0.0284 |
| 12 | 4328521 | rs117233107 | A | G | -0.4705 | 0.1829 | 0.1906 | 0.1344 |
| 12 | 5417856 | rs75507123 | T | G | -0.056 | 0.0632 | -0.1275 | 0.0464 |
| 12 | 8832203 | rs7132012 | A | G | 0.1114 | 0.0441 | 0.1229 | 0.0323 |
| 12 | 13860990 | rs28621435 | A | G | -0.3138 | 0.0729 | -0.1381 | 0.0436 |
| 12 | 15297359 | rs7313556 | A | G | 0.033 | 0.0443 | 0.0973 | 0.0325 |
| 12 | 20754154 | rs73080726 | T | C | -0.1147 | 0.0685 | 0.0425 | 0.0502 |
| 12 | 22015022 | rs704191 | T | C | 0.111 | 0.0428 | -0.047 | 0.0314 |
| 12 | 24210599 | rs7976167 | T | C | 0.1337 | 0.0465 | 0.0345 | 0.034 |
| 12 | 24770878 | rs17287293 | A | G | -0.0369 | 0.0637 | 0.1032 | 0.038 |
| 12 | 26438189 | rs6487543 | A | G | 0.1752 | 0.0563 | 0.1059 | 0.0336 |
| 12 | 27321112 | rs1098708 | A | G | -0.0695 | 0.042 | -0.0855 | 0.0308 |
| 12 | 27962103 | rs10842991 | T | C | -0.2145 | 0.0518 | 0.0016 | 0.0379 |
| 12 | 42540280 | rs7965392 | A | G | 0.1488 | 0.0473 | 0.1334 | 0.0283 |
| 12 | 49981722 | rs7977389 | T | C | 0.1642 | 0.0746 | 0.0799 | 0.0445 |
| 12 | 50537815 | rs7302981 | A | G | 0.3748 | 0.0461 | 0.2306 | 0.0275 |
| 12 | 50767037 | rs61926181 | A | G | -0.3336 | 0.1235 | -0.1357 | 0.0896 |
| 12 | 53440779 | rs73099903 | T | C | 0.4218 | 0.0878 | 0.2185 | 0.0521 |
| 12 | 54443090 | rs7297416 | A | C | 0.2816 | 0.05 | 0.1298 | 0.0299 |
| 12 | 57098040 | rs7137749 | T | C | 0.0294 | 0.0431 | 0.1039 | 0.0317 |
| 12 | 58003922 | rs10437954 | A | G | -0.2398 | 0.0697 | -0.106 | 0.051 |
| 12 | 67782397 | rs4143175 | T | C | 0.3055 | 0.0533 | 0.1375 | 0.0319 |
| 12 | 79685226 | rs7963801 | T | C | -0.1327 | 0.045 | -0.0536 | 0.0329 |
| 12 | 90060586 | rs17249754 | A | G | -0.8015 | 0.0619 | -0.4076 | 0.037 |
| 12 | 94882905 | rs76785029 | T | C | -0.0793 | 0.0973 | 0.1682 | 0.0581 |
| 12 | 95487226 | rs7977311 | T | C | -0.1237 | 0.0668 | 0.0872 | 0.049 |
| 12 | 96109855 | rs11108209 | T | C | -0.206 | 0.0797 | -0.1926 | 0.0476 |
| 12 | 96717095 | rs7134060 | A | G | -0.0603 | 0.0418 | -0.0816 | 0.0306 |
| 12 | 111281636 | rs12184466 | T | C | 0.2956 | 0.0695 | 0.2355 | 0.0413 |
| 12 | 111884608 | rs3184504 | T | C | 0.5774 | 0.0466 | 0.441 | 0.0278 |
| 12 | 115387796 | rs10850411 | T | C | 0.3036 | 0.0494 | 0.1856 | 0.0295 |
| 12 | 115552437 | rs35444 | A | G | 0.3321 | 0.0471 | 0.2201 | 0.0282 |
| 12 | 116198341 | rs11067763 | A | G | 0.1908 | 0.0753 | 0.1429 | 0.045 |
| 12 | 116699675 | rs11615689 | T | C | -0.0826 | 0.0576 | 0.0298 | 0.042 |
| 12 | 120813921 | rs3898618 | T | C | -0.2125 | 0.0952 | -0.2039 | 0.0699 |
| 12 | 122599796 | rs28498002 | T | C | -0.1264 | 0.0489 | -0.1267 | 0.0291 |
| 12 | 123806219 | rs1060105 | T | C | -0.1006 | 0.0572 | -0.1644 | 0.0342 |
| 12 | 124820705 | rs1271309 | A | G | -0.1743 | 0.0581 | -0.1177 | 0.0427 |
| 12 | 133086888 | rs117206641 | T | C | 0.3348 | 0.0783 | 0.2038 | 0.0464 |

| | | | | | | | | |
|---|---|---|---|---|---|---|---|---|
| 13 | 21559858 | rs2480171 | T | C | 0.2057 | 0.0693 | 0.076 | 0.0415 |
| 13 | 22298923 | rs606950 | A | G | 0.1585 | 0.0434 | 0.0723 | 0.0318 |
| 13 | 25257917 | rs55641580 | T | C | 0.1286 | 0.0702 | 0.1268 | 0.0419 |
| 13 | 27115424 | rs1331012 | T | G | 0.1514 | 0.051 | 0.0814 | 0.0304 |
| 13 | 32191408 | rs9532243 | A | C | 0.3595 | 0.0413 | 0.1685 | 0.0303 |
| 13 | 41397482 | rs9549297 | A | G | -0.1049 | 0.0607 | -0.1611 | 0.0362 |
| 13 | 41967193 | rs4274337 | A | G | -0.33 | 0.0612 | -0.1776 | 0.0366 |
| 13 | 42738672 | rs73187288 | A | C | -0.2105 | 0.0712 | -0.0484 | 0.0521 |
| 13 | 47189928 | rs912434 | T | G | 0.1772 | 0.0497 | 0.0144 | 0.0364 |
| 13 | 51489186 | rs9526707 | A | G | -0.1739 | 0.0456 | -0.0613 | 0.0334 |
| 13 | 56398286 | rs75961402 | A | G | 0.2759 | 0.0635 | 0.1297 | 0.038 |
| 13 | 58316637 | rs9563529 | T | G | 0.0554 | 0.0529 | 0.1014 | 0.0386 |
| 13 | 72364382 | rs3861113 | A | C | 0.1821 | 0.0812 | 0.1542 | 0.0487 |
| 13 | 73826901 | rs78474310 | A | G | -0.5088 | 0.1041 | -0.2878 | 0.0762 |
| 13 | 79238925 | rs4304924 | A | G | -0.0696 | 0.0415 | 0.0298 | 0.0305 |
| 13 | 79808655 | rs7988232 | A | G | 0.1392 | 0.0428 | 0.0725 | 0.0313 |
| 13 | 80707408 | rs1215469 | A | C | -0.1972 | 0.0563 | -0.1393 | 0.0336 |
| 13 | 97988689 | rs55684003 | A | G | 0.0602 | 0.0455 | 0.111 | 0.0334 |
| 13 | 111375132 | rs3742182 | T | C | -0.1044 | 0.0524 | 0.0429 | 0.0384 |
| 13 | 113636156 | rs9549328 | T | C | 0.2173 | 0.0553 | 0.0711 | 0.033 |
| 13 | 115000650 | rs7331680 | T | G | 0.3414 | 0.0583 | 0.167 | 0.0427 |
| 14 | 23313633 | rs17880989 | A | G | 0.2086 | 0.1346 | 0.2644 | 0.0979 |
| 14 | 23865885 | rs452036 | A | G | -0.1439 | 0.048 | 0.0863 | 0.0287 |
| 14 | 30122409 | rs17115145 | T | C | 0.1159 | 0.0433 | 0.0401 | 0.0317 |
| 14 | 35110857 | rs4424827 | T | C | -0.1497 | 0.0416 | -0.0849 | 0.0305 |
| 14 | 35871217 | rs8904 | A | G | 0.3201 | 0.0477 | 0.1202 | 0.0285 |
| 14 | 39858442 | rs34983854 | A | G | -0.2259 | 0.0463 | -0.0962 | 0.0276 |
| 14 | 50735947 | rs72683923 | T | C | 1.0239 | 0.1767 | 0.4041 | 0.1311 |
| 14 | 53377540 | rs9888615 | T | C | -0.2356 | 0.0499 | -0.081 | 0.0298 |
| 14 | 54107791 | rs210381 | A | G | -0.2151 | 0.0434 | -0.0093 | 0.0318 |
| 14 | 55285588 | rs7144602 | T | G | -0.1145 | 0.0455 | -0.0219 | 0.0332 |
| 14 | 69260028 | rs57786342 | A | G | 0.1597 | 0.0563 | 0.1082 | 0.0337 |
| 14 | 72462381 | rs11623535 | A | G | 0.1885 | 0.0471 | 0.1363 | 0.0345 |
| 14 | 73279420 | rs4903064 | T | C | -0.039 | 0.0485 | 0.1164 | 0.0356 |
| 14 | 75074316 | rs11159091 | A | G | 0.1433 | 0.0422 | 0.019 | 0.0309 |
| 14 | 89565130 | rs4904503 | T | C | 0.141 | 0.0498 | -0.0391 | 0.0297 |
| 14 | 93112102 | rs11160085 | T | C | 0.1327 | 0.052 | 0.0144 | 0.031 |
| 14 | 94465789 | rs8013933 | T | C | 0.0513 | 0.0467 | -0.0563 | 0.0342 |
| 14 | 98587630 | rs9323988 | T | C | -0.2377 | 0.0464 | -0.1105 | 0.0277 |
| 14 | 100225144 | rs1475130 | T | C | -0.1638 | 0.0485 | 1.00E-04 | 0.029 |
| 14 | 100742658 | rs28470843 | T | C | 0.0482 | 0.043 | -0.0414 | 0.0315 |
| 14 | 103859962 | rs8014182 | T | C | -0.3509 | 0.062 | -0.1593 | 0.0452 |
| 14 | 104620193 | rs34161718 | T | C | -0.0417 | 0.0614 | -0.1378 | 0.0367 |
| 15 | 26105602 | rs10873612 | T | C | -0.0995 | 0.0476 | -0.1062 | 0.0285 |
| 15 | 40317075 | rs11629850 | A | G | 0.1417 | 0.0416 | 0.1053 | 0.0305 |
| 15 | 41311799 | rs2925345 | T | C | 0.1559 | 0.0419 | 0.0706 | 0.0307 |
| 15 | 41974660 | rs4924570 | T | C | 0.0218 | 0.0461 | -0.1047 | 0.0337 |
| 15 | 48914926 | rs1036477 | A | G | 0.3401 | 0.0739 | -0.0656 | 0.0442 |
| 15 | 50810621 | rs3098186 | T | C | -0.1515 | 0.0428 | -0.0246 | 0.0313 |
| 15 | 59429160 | rs3191402 | A | G | -0.1832 | 0.0463 | -0.0176 | 0.0339 |
| 15 | 62808539 | rs956006 | T | C | -0.0698 | 0.0493 | 0.0425 | 0.0295 |
| 15 | 65166309 | rs832890 | T | C | 0.0691 | 0.0418 | -0.0471 | 0.0307 |
| 15 | 66869072 | rs7178615 | A | G | -0.1414 | 0.0473 | -0.1328 | 0.0283 |
| 15 | 68454523 | rs62004794 | A | G | -0.0166 | 0.0419 | -0.0905 | 0.0307 |
| 15 | 71621524 | rs11853359 | A | G | 0.048 | 0.0443 | -0.1445 | 0.0325 |
| 15 | 74557817 | rs61653296 | A | G | -0.1387 | 0.0572 | -0.1667 | 0.0341 |
| 15 | 75077367 | rs1378942 | A | C | -0.4867 | 0.0481 | -0.3725 | 0.0288 |
| 15 | 79156983 | rs62011052 | T | C | -0.0165 | 0.0644 | 0.1532 | 0.0385 |
| 15 | 81016227 | rs2759308 | A | G | 0.2592 | 0.046 | 0.1176 | 0.0275 |
| 15 | 83799632 | rs2034618 | T | C | -0.0151 | 0.0549 | -0.1093 | 0.0329 |
| 15 | 85162551 | rs7180952 | T | C | -0.0125 | 0.042 | -0.0911 | 0.0308 |
| 15 | 85680532 | rs3743157 | A | C | 0.1997 | 0.0549 | 0.0673 | 0.0403 |
| 15 | 90641809 | rs28611491 | T | C | 0.1289 | 0.0732 | -0.0679 | 0.0535 |
| 15 | 95312071 | rs12906962 | T | C | -0.2663 | 0.0493 | -0.1883 | 0.0295 |
| 15 | 96635898 | rs4984496 | T | G | 0.0718 | 0.0451 | 0.1024 | 0.0331 |
| 16 | 706067 | rs9932866 | A | G | 0.104 | 0.0425 | 0.0935 | 0.0311 |
| 16 | 1344291 | rs11248862 | A | G | 0.101 | 0.0657 | -0.0584 | 0.048 |

| | | | | | | | | |
|---|---|---|---|---|---|---|---|---|
| 16 | 4297651 | rs4785955 | T | G | 0.1369 | 0.0574 | -0.0248 | 0.0341 |
| 16 | 4943019 | rs12921187 | T | G | -0.2421 | 0.0458 | -0.1561 | 0.0274 |
| 16 | 6889675 | rs35450617 | T | G | -0.1298 | 0.0458 | -0.0548 | 0.0335 |
| 16 | 11198835 | rs11642631 | T | C | -0.0504 | 0.0421 | 0.0507 | 0.0308 |
| 16 | 14487036 | rs57327054 | T | C | -0.0831 | 0.0509 | -0.0951 | 0.0305 |
| 16 | 15912544 | rs3915425 | T | C | 0.1358 | 0.045 | 0.0174 | 0.033 |
| 16 | 20365654 | rs13333226 | A | G | 0.336 | 0.058 | 0.253 | 0.0347 |
| 16 | 30111904 | rs6565174 | A | C | -0.2231 | 0.068 | -0.1404 | 0.05 |
| 16 | 30936743 | rs72799341 | A | G | 0.0949 | 0.054 | 0.1572 | 0.0323 |
| 16 | 49768046 | rs10468291 | A | C | -0.1175 | 0.0463 | -0.1233 | 0.0277 |
| 16 | 50550137 | rs34941092 | A | G | -0.302 | 0.0651 | -0.1933 | 0.0388 |
| 16 | 51758116 | rs9932220 | A | G | -0.2235 | 0.0549 | -0.1714 | 0.0328 |
| 16 | 58566304 | rs37060 | A | G | 0.1201 | 0.047 | -0.0192 | 0.0344 |
| 16 | 65282820 | rs28633979 | A | C | -0.0832 | 0.042 | 0.0349 | 0.0308 |
| 16 | 66914492 | rs45474499 | T | C | 0.2763 | 0.101 | 0.3021 | 0.0741 |
| 16 | 69640217 | rs33063 | A | G | 0.1777 | 0.0647 | -0.0689 | 0.0386 |
| 16 | 75444572 | rs35261357 | T | C | 0.2778 | 0.0467 | 0.1025 | 0.0279 |
| 16 | 80864776 | rs56844452 | T | C | -0.3427 | 0.0911 | -0.0138 | 0.0545 |
| 16 | 81574197 | rs8059962 | T | C | -0.1688 | 0.0467 | -0.1138 | 0.0279 |
| 16 | 83045790 | rs7500448 | A | G | 0.225 | 0.0533 | -0.0262 | 0.0319 |
| 16 | 85318302 | rs7187540 | A | C | -0.1431 | 0.0458 | -0.0579 | 0.0335 |
| 16 | 87993889 | rs6540125 | T | G | 0.1864 | 0.0475 | 0.0714 | 0.0284 |
| 17 | 1333598 | rs12941318 | T | C | -0.208 | 0.0484 | -0.0798 | 0.0288 |
| 17 | 1958609 | rs4480845 | T | C | 0.1473 | 0.0447 | -0.0067 | 0.0327 |
| 17 | 3880148 | rs7215084 | T | C | 0.1662 | 0.0416 | 0.1443 | 0.0305 |
| 17 | 6473882 | rs28427409 | T | C | 0.1641 | 0.0463 | -0.0026 | 0.0277 |
| 17 | 7571752 | rs78378222 | T | G | 0.1831 | 0.2082 | -0.6128 | 0.1238 |
| 17 | 8078765 | rs8069739 | T | C | -0.1594 | 0.0441 | -0.137 | 0.0324 |
| 17 | 18185510 | rs4925159 | A | G | 0.2482 | 0.0419 | 0.2029 | 0.0307 |
| 17 | 27195674 | rs138285687 | T | C | -0.396 | 0.1098 | 0.0023 | 0.0807 |
| 17 | 29161503 | rs11080134 | A | G | -0.0927 | 0.0432 | -0.1031 | 0.0317 |
| 17 | 30032420 | rs1551355 | T | C | 0.1618 | 0.049 | 0.1193 | 0.0359 |
| 17 | 33313729 | rs3135967 | A | G | 0.0987 | 0.0422 | -0.0415 | 0.0309 |
| 17 | 40317241 | rs79089478 | T | C | 0.1399 | 0.1461 | -0.0656 | 0.087 |
| 17 | 40919596 | rs56228409 | A | C | -0.0848 | 0.0571 | 0.0685 | 0.0418 |
| 17 | 42680402 | rs9904409 | A | G | 0.2069 | 0.0707 | 0.0196 | 0.0519 |
| 17 | 45013271 | rs17608766 | T | C | -0.6153 | 0.0669 | -0.1889 | 0.0399 |
| 17 | 45888374 | rs62076103 | A | G | -0.26 | 0.0916 | 0.071 | 0.0672 |
| 17 | 46688256 | rs7406910 | T | C | -0.4877 | 0.0812 | -0.1593 | 0.0485 |
| 17 | 47402807 | rs12940887 | T | C | 0.2657 | 0.0469 | 0.2476 | 0.028 |
| 17 | 57853214 | rs2645466 | A | C | -0.1964 | 0.0494 | -0.0304 | 0.0295 |
| 17 | 58950791 | rs1036902 | T | C | -0.1786 | 0.0566 | -0.0597 | 0.0415 |
| 17 | 59485393 | rs2240736 | T | C | 0.4265 | 0.0525 | 0.2483 | 0.0314 |
| 17 | 60767151 | rs740698 | T | C | -0.1458 | 0.0477 | 0.0163 | 0.0285 |
| 17 | 61559625 | rs4308 | A | G | 0.2728 | 0.0477 | 0.1832 | 0.0285 |
| 17 | 62381714 | rs6504213 | T | C | -0.1381 | 0.0437 | 0.0108 | 0.032 |
| 17 | 64252393 | rs112260610 | T | C | 0.3389 | 0.0669 | 0.1428 | 0.0398 |
| 17 | 73949045 | rs2467099 | T | C | -0.191 | 0.0547 | -0.0837 | 0.0329 |
| 17 | 74686809 | rs35504735 | A | G | 0.0612 | 0.0415 | -0.0629 | 0.0304 |
| 17 | 76799898 | rs9302885 | A | G | 0.1367 | 0.0421 | 0.028 | 0.0309 |
| 17 | 79367409 | rs112280096 | A | C | -0.1169 | 0.0431 | -0.0781 | 0.0317 |
| 18 | 12711052 | rs963920 | T | G | -0.1135 | 0.0458 | 0.0228 | 0.0336 |
| 18 | 20158965 | rs4800420 | A | G | -0.0381 | 0.0451 | 0.0884 | 0.033 |
| 18 | 24546824 | rs1154214 | T | G | -0.2163 | 0.046 | -0.0862 | 0.0275 |
| 18 | 31161426 | rs10164193 | T | G | -0.3638 | 0.0854 | -0.2828 | 0.0511 |
| 18 | 34289285 | rs61735998 | T | G | 0.2049 | 0.136 | -0.166 | 0.0997 |
| 18 | 42141977 | rs12958173 | A | C | 0.3518 | 0.0495 | 0.1774 | 0.0296 |
| 18 | 43097750 | rs7236548 | A | C | 0.2643 | 0.0582 | 0.0047 | 0.0348 |
| 18 | 48142854 | rs745821 | T | G | 0.1772 | 0.0526 | 0.1411 | 0.0315 |
| 18 | 48283949 | rs36010659 | T | C | 0.195 | 0.0657 | -0.0047 | 0.0392 |
| 18 | 48799991 | rs11876341 | A | G | -0.2406 | 0.0461 | -0.1213 | 0.0338 |
| 18 | 51851616 | rs34163044 | A | C | 0.1036 | 0.0441 | 0.1494 | 0.0323 |
| 18 | 52607301 | rs72930904 | T | C | -0.2255 | 0.0622 | -0.1535 | 0.0372 |
| 18 | 54578482 | rs10048404 | T | C | -0.1412 | 0.0513 | -0.0586 | 0.0374 |
| 18 | 55732115 | rs7235890 | T | G | -0.2597 | 0.0757 | -0.1875 | 0.0453 |
| 18 | 57829135 | rs6567160 | T | C | 0.1618 | 0.0541 | 0.0847 | 0.0324 |
| 18 | 60223017 | rs12172847 | A | G | -0.1532 | 0.0453 | -0.0582 | 0.0332 |

| | | | | | | | | |
|---|---|---|---|---|---|---|---|---|
| 18 | 60845884 | rs12454712 | T | C | 0.2258 | 0.0426 | 0.0902 | 0.0312 |
| 18 | 73034151 | rs10460108 | A | G | 0.2039 | 0.0452 | 0.115 | 0.027 |
| 18 | 74070562 | rs1047922 | T | C | -0.1594 | 0.0711 | 0.0246 | 0.0425 |
| 19 | 670234 | rs7250835 | T | C | 0.1644 | 0.0596 | -0.0512 | 0.0436 |
| 19 | 1435771 | rs3760994 | A | G | -0.127 | 0.0531 | -0.0422 | 0.0314 |
| 19 | 2232221 | rs740406 | A | G | -0.5488 | 0.1017 | -0.1985 | 0.0603 |
| 19 | 5066330 | rs2613765 | A | G | -0.2304 | 0.0415 | -0.0594 | 0.0304 |
| 19 | 7224431 | rs7248104 | A | G | -0.1589 | 0.0463 | -0.0402 | 0.0277 |
| 19 | 7252756 | rs4247374 | T | C | -0.5063 | 0.0753 | -0.3389 | 0.045 |
| 19 | 8398714 | rs2009733 | A | G | 0.1064 | 0.0475 | 0.0983 | 0.0283 |
| 19 | 10332988 | rs10409243 | T | C | -0.1088 | 0.0443 | -0.0914 | 0.0324 |
| 19 | 10841472 | rs1529744 | T | C | -0.0138 | 0.0453 | 0.0926 | 0.0332 |
| 19 | 11526765 | rs167479 | T | G | -0.4284 | 0.0562 | -0.3415 | 0.034 |
| 19 | 11584818 | rs17638167 | T | C | -0.5228 | 0.1095 | -0.2379 | 0.0656 |
| 19 | 16436262 | rs3745318 | T | C | 0.2041 | 0.0565 | 0.1439 | 0.0335 |
| 19 | 17222584 | rs1077795 | A | G | 0.1486 | 0.0518 | 0.0961 | 0.0378 |
| 19 | 18558876 | rs8111708 | A | G | 0.0359 | 0.0479 | -0.0657 | 0.0286 |
| 19 | 19789528 | rs2304130 | A | G | -0.3072 | 0.086 | -0.2336 | 0.0512 |
| 19 | 21950402 | rs6511291 | T | C | -0.0953 | 0.0472 | -0.0757 | 0.0281 |
| 19 | 30294991 | rs62104477 | T | G | 0.087 | 0.0487 | 0.1301 | 0.0291 |
| 19 | 31927547 | rs8105753 | A | C | 0.1895 | 0.0487 | 0.0917 | 0.0291 |
| 19 | 32590773 | rs1821295 | T | C | -0.1021 | 0.0457 | -0.1243 | 0.0335 |
| 19 | 33889593 | rs7256564 | A | G | 0.2039 | 0.0487 | 0.1093 | 0.0291 |
| 19 | 39438532 | rs12983238 | A | G | -0.1717 | 0.0575 | -0.1589 | 0.0342 |
| 19 | 41858921 | rs1800470 | A | G | -0.0721 | 0.0426 | 0.0363 | 0.0312 |
| 19 | 45412079 | rs7412 | T | C | -0.3363 | 0.0753 | 0.0591 | 0.055 |
| 19 | 46180414 | rs34783010 | T | G | 0.1463 | 0.0506 | 0.0137 | 0.0371 |
| 19 | 49605705 | rs73046792 | A | G | -0.2413 | 0.069 | -0.1067 | 0.0412 |
| 19 | 50935809 | rs138877676 | T | G | -0.6453 | 0.1882 | -0.4843 | 0.1374 |
| 20 | 2793063 | rs2143635 | T | C | -0.1816 | 0.0793 | 0.0341 | 0.0474 |
| 20 | 6657554 | rs11087740 | T | C | -0.0985 | 0.0417 | 0.0527 | 0.0306 |
| 20 | 8626271 | rs6108168 | A | C | -0.2037 | 0.0519 | -0.1415 | 0.031 |
| 20 | 10458688 | rs680515 | A | G | -0.1031 | 0.0447 | 0.113 | 0.0327 |
| 20 | 10969030 | rs1327235 | A | G | -0.3548 | 0.045 | -0.239 | 0.0269 |
| 20 | 11886643 | rs1232482 | T | C | -0.1187 | 0.0424 | -0.0969 | 0.0311 |
| 20 | 17882452 | rs2618647 | A | G | -0.1773 | 0.0455 | -0.1199 | 0.0272 |
| 20 | 19465907 | rs6081613 | A | G | 0.1358 | 0.0506 | -0.088 | 0.0302 |
| 20 | 30169673 | rs6060114 | T | C | 0.2809 | 0.062 | 0.1617 | 0.037 |
| 20 | 32298286 | rs13042148 | T | C | -0.2515 | 0.0657 | -0.1878 | 0.0393 |
| 20 | 36849007 | rs4811601 | T | C | -0.0511 | 0.0422 | -0.1423 | 0.0309 |
| 20 | 42797358 | rs6031435 | A | G | -0.2268 | 0.0456 | -0.1111 | 0.0273 |
| 20 | 47308798 | rs6095241 | A | G | -0.1884 | 0.0454 | -0.1238 | 0.0271 |
| 20 | 48004238 | rs237485 | A | G | 0.1551 | 0.0505 | 0.1157 | 0.0301 |
| 20 | 50108980 | rs6021247 | A | G | 0.1506 | 0.0411 | 0.0912 | 0.0302 |
| 20 | 57751117 | rs6015450 | A | G | -0.655 | 0.0691 | -0.4801 | 0.0413 |
| 20 | 62694319 | rs35213536 | T | G | 0.3396 | 0.0531 | 0.1313 | 0.0388 |
| 21 | 16556367 | rs1882961 | T | C | 0.2147 | 0.0466 | 0.1033 | 0.0341 |
| 21 | 35596842 | rs9976596 | T | C | 0.0785 | 0.0599 | -0.0832 | 0.0438 |
| 21 | 37692507 | rs62229372 | T | C | 0.2217 | 0.072 | 0.1568 | 0.0523 |
| 21 | 44760603 | rs12627651 | A | G | 0.4162 | 0.0528 | 0.2479 | 0.0316 |
| 21 | 45107562 | rs9306160 | T | C | -0.2069 | 0.0474 | -0.1687 | 0.0284 |
| 21 | 47422412 | rs35796750 | T | C | -0.1108 | 0.0467 | 0.0091 | 0.0278 |
| 21 | 47962811 | rs11701512 | A | G | 0.0645 | 0.0552 | -0.0517 | 0.0403 |
| 22 | 19967980 | rs12628032 | T | C | 0.1972 | 0.0499 | 0.018 | 0.0298 |
| 22 | 28056338 | rs134041 | T | C | -0.0576 | 0.0432 | 0.079 | 0.0317 |
| 22 | 28921347 | rs9608690 | A | G | -0.308 | 0.0912 | -0.1563 | 0.0544 |
| 22 | 29451671 | rs4823006 | A | G | 0.1407 | 0.0455 | 0.1098 | 0.0272 |
| 22 | 30768777 | rs5753103 | A | G | 0.1031 | 0.0459 | -0.0245 | 0.0274 |
| 22 | 32517431 | rs9609429 | T | C | 0.1626 | 0.0513 | 0.1423 | 0.0307 |
| 22 | 38117943 | rs5750482 | T | C | 0.1127 | 0.0429 | 0.0076 | 0.0314 |
| 22 | 40729614 | rs470113 | A | G | -0.0784 | 0.058 | 0.0599 | 0.0346 |
| 22 | 42038786 | rs73161324 | T | C | 0.2121 | 0.1156 | -0.1117 | 0.0689 |
| 22 | 50219952 | rs77692990 | T | C | -0.1628 | 0.076 | -0.0047 | 0.0557 |
| 22 | 50727921 | rs28578714 | T | C | 0.1761 | 0.0421 | 0.1008 | 0.0309 |

**Supplemental table 2. Description of each of 5 cognitive tests administered to UK Biobank participants.**

| Cognitive test | Description |
|---|---|
| **Pairs Memory test** | Participants are asked to memorize the position of as many matching pairs of cards as possible. The cards are then turned face down on the screen and the participant is asked to touch as many pairs as possible in the fewest tries. Multiple rounds were conducted. The first round used 3 pairs of cards and the second 6 pairs of cards. The result of the test is the total number of incorrect matches. |
| **Reaction Time test** | Based on 12 rounds of the card-game 'Snap'. In each round, the participant is shown two cards at a time; if both cards are the same, they press a button-box that is on the table in front of them as quickly as possible. The result of the test is the mean time it took to first click the 'snap' button across all 12 rounds. |
| **Numeric Memory test** | This test designed to assess numeric short-term memory, as part of the touchscreen questionnaire. The participant was shown a 2-digit number to remember. The number then disappeared and after a short while they were asked to enter the number onto the screen. The number became one digit longer each time they remembered correctly (up to a maximum of 12 digits). The result of the test is the number of items remembered correctly. |
| **Prospective Memory test** | Early in the touchscreen cognitive section, the participant is shown the message "At the end of the games we will show you four colored shapes and ask you to touch the Blue Square. However, to test your memory, we want you to actually touch the Orange Circle instead." The result of this test is whether the task was completed correctly. |
| **Fluid Intelligence test** | This test evaluates the capacity to solve problems that require logic and reasoning ability, independent of acquired knowledge. The participant has 2 minutes to complete as many questions as possible from the test. The result of this test is the number of questions answered correctly. |

**Supplemental Table 3. Population characteristics for participants of all race/ethnic groups who completed all five cognitive tests at baseline.**

| Variable, n (%) | All Study participants<br><br>n=48,118 | Low polygenic risk for hypertension*<br><br>n=9,625 | Intermediate polygenic risk for hypertension*<br><br>n=28,8870 | High polygenic risk for hypertension*<br><br>n=9,623 | P value<br><br>Comparison across genetic risk categories |
|---|---|---|---|---|---|
| **Demographics** | | | | | |
| **Age** | 56.4 (8.3) | 56.4 (8.3) | 56.4 (8.3) | 56.5 (8.3) | 0.51 |
| **Male sex** | 2,1921 (45.6) | 4,429 (46.1) | 13,157 (45.7) | 4,294 (44.7) | 0.13 |
| **European** | 42,080 (87.5) | 9,118 (94.9) | 27,542 (95.4) | 9,323 (96.8) | <0.001 |
| **Black** | 332 (0.7) | 60 (0.6) | 213 (0.8) | 59 (0.7) | |
| **Asian** | 976 (2.0) | 275 (2.9) | 584 (2.0) | 114 (1.2) | |
| **Other race/ethnicity** | 4,730 (9.8) | 172 (1.6) | 531 (1.8) | 127 (1.3) | |
| **Vascular risk factors** | | | | | |
| **Hypertension** | 14,133 (29.4) | 2,158 (22.5) | 8,392 (29.1) | 3,565 (37.1) | <0.001 |
| **Hyperlipidemia** | 6,638 (13.8) | 1,188 (12.4) | 4,007 (13.9) | 1,434 (14.9) | <0.001 |
| **Diabetes** | 2,354 (4.9) | 446 (4.6) | 1,398 (4.9) | 507 (5.3) | 0.11 |
| **Current smoker** | 4,758 (9.9) | 942 (9.8) | 2,880 (10.0) | 927 (9.7) | 0.65 |
| **Prior smoker** | 16,941 (35.3) | 3,339 (34.9) | 10,153 (35.3) | 3,415 (35.7) | |
| **BMI, mean (SD)** | 27.36 (4.75) | 27.47 (4.79) | 27.34 (4.77) | 27.28 (4.64) | 0.014 |
| **Comorbidities** | | | | | |
| **Myocardial infarction** | 1417 (2.9) | 227 (2.4) | 863 (3.0) | 323 (3.4) | <0.001 |
| **Atrial fibrillation** | 1999 (4.2) | 377 (3.9) | 1185 (4.1) | 433 (4.5) | 0.110 |
| **Admission BP, mean (SD)** | | | | | |
| **Systolic BP** | 138.8 (18.8) | 134.4 (17.9) | 138.8 (18.6) | 143.2 (19.4) | <0.001 |
| **Diastolic BP** | 82.44 (10.03) | 80.68 (9.84) | 82.46 (9.96) | 84.17 (10.15) | <0.001 |

Abbreviations: BMI = body mass index; SD = standard deviation; BP = blood pressure.

* Polygenic risk for hypertension estimated using genetic risk variants for systolic blood pressure.

**Supplemental Table 4. Association between polygenic susceptibility to hypertension and baseline cognitive status in participants of all race/ethnic groups.**

| Linear regression model | Comparison of polygenic susceptibility to hypertension | Polygenic susceptibility to hypertension Genetic risk variants for systolic BP | | | Polygenic susceptibility to hypertension Genetic risk variants for diastolic BP | | |
|---|---|---|---|---|---|---|---|
| | | % change | Beta (SE) | Test for trend p | % change | Beta (SE) | Test for trend p |
| **Model 1 Adjusted for age, sex and PC 1 through 4** | Low versus intermediate genetic risk | -2.7% | -0.027 (0.011) | <0.001 | -1.9% | -0.019 (0.011) | 0.03 |
| | Low versus high genetic risk | -5.0% | -0.050 (0.014) | | -3.0% | -0.030 (0.014) | |
| **Model 2 Unadjusted** | Low versus intermediate genetic risk | -2.1% | -0.021 (0.012) | 0.02 | -2.1% | -0.021 (0.012) | 0.19 |
| | Low versus high genetic risk | -3.4% | -0.034 (0.014) | | -1.9% | -0.019 (0.014) | |
| **Model 3 Adjusted for age, sex, diabetes, hyperlipidemia, smoking and PC 1 through 4** | Low versus intermediate genetic risk | -2.3% | -0.023 (0.011) | <0.001 | -1.6% | -0.016 (0.011) | 0.06 |
| | Low versus high genetic risk | -4.4% | -0.044 (0.014) | | -2.5% | -0.025 (0.014) | |
| **Model 4 Adjusted for age, sex, diabetes, hyperlipidemia, smoking, history of myocardial infarction and PC 1 through 4** | Low versus intermediate genetic risk | -2.3% | -0.023 (0.011) | 0.002 | -1.5% | -0.015 (0.011) | 0.07 |
| | Low versus high genetic risk | -4.3% | -0.043 (0.014) | | -2.4% | -0.024 (0.014) | |

**Supplemental Table 5. Association between polygenic susceptibility to hypertension and results of 5 different cognitive tests in participants of all race/ethnic groups.**

| Cognitive test | Analytical approach | Test interpretation | Comparison of polygenic susceptibility to hypertension | Polygenic susceptibility to hypertension Genetic risk variants for systolic BP | | Polygenic susceptibility to hypertension Genetic risk variants for diastolic BP | |
|---|---|---|---|---|---|---|---|
| | | | | Beta (SE) | Test for trend p | Beta (SE) | Test for trend p |
| **Pairs Memory test** | Linear regression | Lower values imply better cognitive status | Low versus intermediate genetic risk | 0.008 (0.011) | 0.42 | -0.003 (0.011) | 0.92 |
| | | | Low versus high genetic risk | 0.011 (0.013) | | 0.001 (0.013) | |
| **Reaction Time test** | Linear regression | Lower values imply better cognitive status | Low versus intermediate genetic risk | 0.015 (0.011) | 0.11 | 0.012 (0.011) | 0.03 |
| | | | Low versus high genetic risk | 0.023 (0.014) | | 0.030 (0.014) | |
| **Numeric Memory test** | Linear regression | Higher values imply better cognitive status | Low versus intermediate genetic risk | -0.019 (0.011) | 0.02 | -0.011 (0.011) | 0.40 |
| | | | Low versus high genetic risk | -0.030 (0.013) | | -0.011 (0.013) | |
| **Prospective Memory test** | Logistic regression | Higher values imply better cognitive status | Low versus intermediate genetic risk | -0.014 (0.029) | 0.35 | 0.012 (0.029) | 0.72 |
| | | | Low versus high genetic risk | -0.033 (0.036) | | -0.013 (0.036) | |
| **Fluid Intelligence test** | Linear regression | Higher values imply better cognitive status | Low versus intermediate genetic risk | -0.023 (0.011) | 0.0002 | -0.023 (0.011) | 0.06 |
| | | | Low versus high genetic risk | -0.052 (0.014) | | -0.026 (0.014) | |
